# Estimating the Global Spread of Epidemic Human Monkeypox with Bayesian Directed Acyclic Graphic Model

**DOI:** 10.1101/2022.12.16.22283570

**Authors:** Ling-Chun Liao, Chen-Yang Hsu, Chao-Chih Lai, Tony Hsiu-Hsi Chen

## Abstract

**Objectives:** A ‘Public Health Emergency of International Concern (PHEIC)’ monkeypox outbreak was declared by the World Health Organization on June 23, 2022. More than 16,000 monkeypox cases were reported in more than 75 countries across six regions as July 25.

**Design:** A modeling study.

**Setting and Participants:** Daily confirmed Monkeypox cases of the global, United States, Spain, Brazil, and United Kingdom were retrieved from the Global health team till September 23,2022. All conducted analyses are at the aggregate level.

without involvement of confidential information.

**Primary and secondary outcome measures:** The Bayesian SIR (*Susceptible-Infected-Recovered*) model with Directed Acyclic Graphic method was used to estimate the basic/effective reproductive number (R_0_/ R_e_) and to assess the epidemic spread of Monkeypox in the globe.

**Results:** The maximum estimated R_0_/ R_e_ was 1.16 (1.15-1.17), 1.20 (1.20-1.20), 1.34 (1.34-1.35), and 1.33 (1.33-1.33) in United States, Spain, Brazil, and United Kingdom, respectively. The values of R_0_/ R_e_ was toward to below 1 after August, 2022. The estimated infectious time before isolation ranged from 2.05 to 2.74 days.

**Conclusions:** The PHEIC of global spreading of human Monkeypox outside Africa has been contained so as to avoid a pandemic in the light of the reasoning-based epidemic model assessment.

**Strengths and limitations of this study:**

- We estimated the epidemiology parameters based on the Bayesian SIR model for the uncertainty transmission of monkeypox under assumptions of homogeneous random mixing population and surveillance systems were the same across countries.
- This approach can minimize biases between population, different surveillance systems across countries. Estimating results of R0 were limited but consistent between countries and similar to the simulation data by the deterministic SIR model.

## Introduction

Monkeypox is a rare zoonotic disease, and the virus was initially discovered in 1958. (1) The first human case with monkeypox, which is caused by the monkeypox virus, was diagnosed in 1970. (2) Monkeypox mainly occurs in forested rural areas in central and Western Africa. Most cases acquired with the Central African clade and the others with the West African clade from 1970 to 2019. (3) Since the cessation of smallpox vaccination, the incidence of reported monkeypox cases, the median age of patients, and outbreaks are rising. In African, about 72.5% monkeypox cases were suspected via the animal to human transmission, and 27.5% cases were by a human source in 1980s. (4) But 78% monkeypox cases were infected by a human source in 1990s. (5) However, most cases outside of African were suspected via the animal to human transmission. (3) Therefore, human-to-human transmission was inefficient because sporadic outbreaks and travel associated cases outside Africa had limited secondary spread before April, 2022. Unfortunately, more than 16,000 monkeypox cases were reported in more than 75 countries across six regions as July 25, 2022, (6) and the majority of cases (98%) have been confirmed since early May 2022. In addition, the World Health Organization to declare a “Public Health Emergency of International Concern (PHEIC)” monkeypox outbreak on June 23, 2022. (6) Over 7,000 monkeypox cases has been confirmed in the United States as August 5, 2022. (7) Over 90% of monkeypox cases have occurred mainly among men through sexual contexts. (8, 9) Several reasons including waning smallpox immunity, relaxing COVID-19 contain measures, and sexual activities after large gathering were linked to the global outbreak of monkeypox virus infection. (8) Avoiding human contact by good hygiene (10) and early detection (9), reducing the possibility of transmission by pre-exposure and post-exposure vaccination and treatment (11) are the major contain measures to prevent the outbreak of monkeypox. Thus, the surveillance of the spread of monkeypox is important to know the effectiveness of these contain measures.

The basic/effective reproductive number (R_0_/R_e_, the expected number of secondary infections) is a useful indicator for the spread of Monkeypox. Aim of this study is to estimate R_0_/R_e_ of Monkeypox by the Bayesian SIR (*Susceptible-Infected-Recovered*) model to elucidate the spread of Monkeypox. It is indispensable to determine whether or not Monkeypox will continually spread through a population.

## Materials

The data on the Monkeypox outbreaks was retrieved from the Global health team for estimating of R_0_/R_e_. (8, 9) The data of the global, United States, Spain, Brazil, and United Kingdom retrieved till September 23,2022.

## Methods

The Bayesian ordinary differential equation (ODE) SIR model, including S (susceptible population), I (infectious persons with Monkeypox), and R (recovered persons from infectious status or infectious persons being isolated) states, was used to estimate the basic reproductive number and to predict the global propagation of Monkeypox outbreak making use of the observed number of cases with the consideration of uncertainty. (10) Let s(t), i(t), and r(t) denote the numbers of susceptible, infectives, and recovered/isolated at time t. It can be depicted by using the ordinary differential equations as follow:

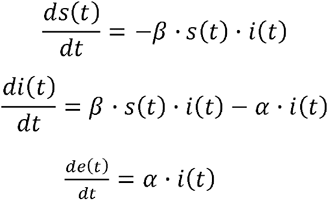

The parameters of transmission coefficient (β) and recovery rate (α) from the infectious status can be estimated. Note that the population of susceptible is fixed, the following equation always holds. s(t)+i(t)+r(t)=N. Two random variables, number of unknown infected status [s(t)] and the confirmed cases per day [i(t)+r(t)] following two distributions of

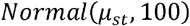

and

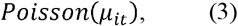

respectively. The basic reproduction number (R_0_) were estimated as 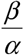. The informative priors for the parameters a was used to fit the mean duration from infectious to recovered for 8.5 days (95% CI: 6.6-10.9) [a∼ gamma (26.7, inverse-scale=222.92)] according to the incubation period of monkeypox in the literature. (11)

The Bayesian directed acyclic graphic (DAG) model for estimating the spread of human moneypox 3-state model is illustrated in Figure 1. By using the equation derived above, we can link the parameters of transmission coefficient (β) and recovery rate (α) from the infectious status with the expected number of susceptible and infective cases at day *t*. The latter two components were used as the scale parameters of the normal distribution for the observed node of susceptible and of the Poisson distribution for the observed node of infective cases. R_0_/R_e_ can be derived with a logic function to transmission coefficient and recovery rate. The required smallpox vaccine coverage for containing mokeypox given an 85% efficacy (16) can be further derived. As far as the stochastic link is concerned, the random variable of *s*_*t*_ follows a normal distribution indicated above. The corresponding distribution for the random variable of *i*_*t*_ is Poisson distribution.

**Figure 1.**
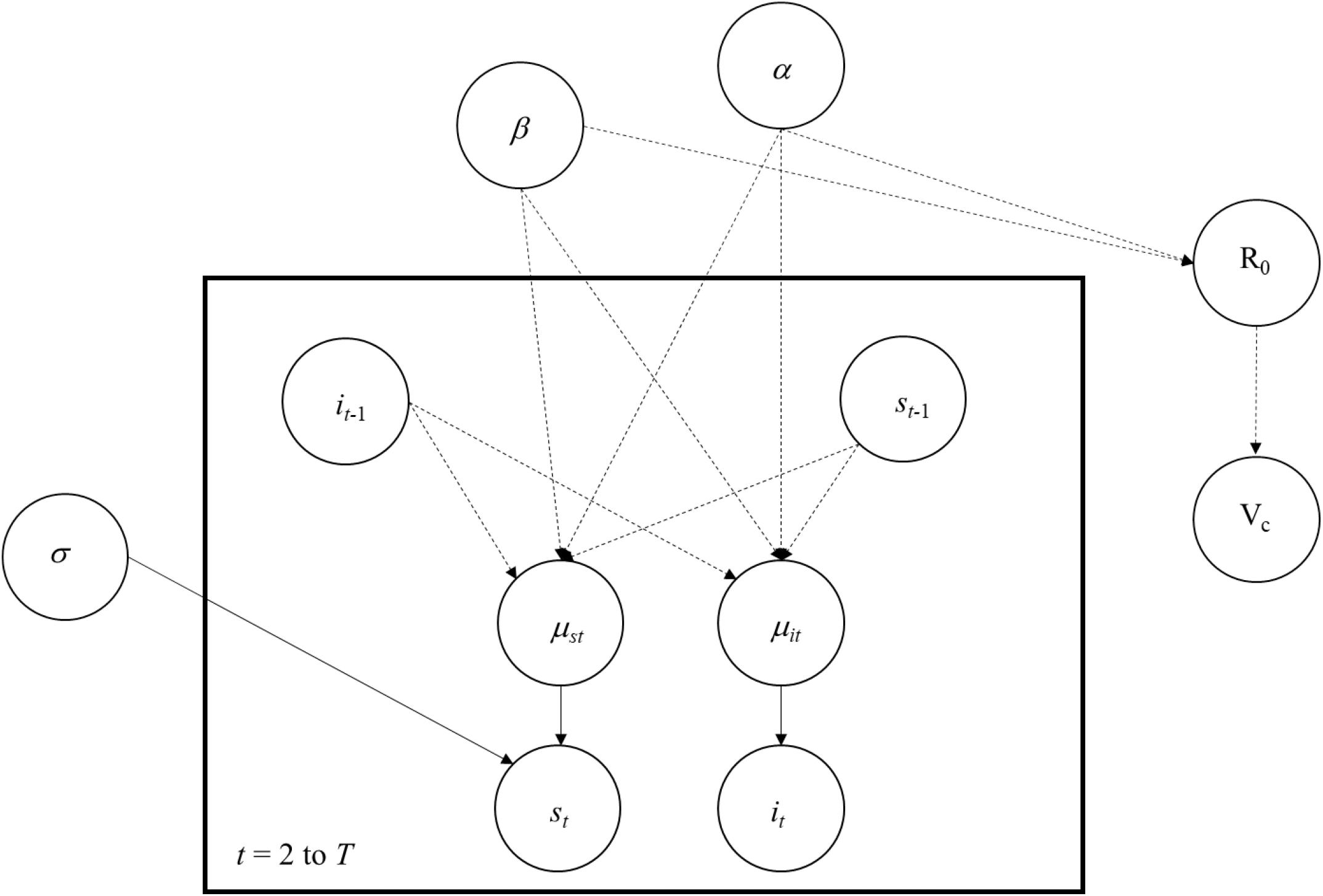
The directed acyclic graph model for estimating the spread of human moneypox. Annotation: *T*: total days of observation. *st* and *i*_*t*_: number of susceptible and infective subjects at day *t*. *µ*_*st*_ and *µ*_*it*_: expected number of subjects susceptible and infective subjects at day *t*. *α* and *β*: the parameters of transmission coefficient (β) and recovery rate (α) from the infectious status *R*_0_/R_e_: the basic/effective reproduction number V_c_: the minimum vaccine coverage in order to control the disease Dashed arrow and solid arrow refer to logit link and stochastic link, respectively.

### Patient and public involvement

Patients and/or the public were not involved in the design, or conduct, or reporting, or dissemination plans of this research.

## Results

The parameters estimated by the Bayesian SIR model from the Monkeypox cases in United States, Spain, Brazil, United Kingdom and on the globe are listed in Table 1. The estimated basic reproductive number (R0) or effective reproductive number (Re) of Monkeypox ranged from 1.00 (0.99-1.15) to 1.46 (1.37-1.51) on the global. The estimated required smallpox vaccine coverage for containing the spread of monkeypox was highest in May 2022 (37%). The maximum estimated R0/ Re was 1.20 (1.197-1.201), 1.19 (1.19-1.19), 1.34 (1.34-1.35), and 1.33 (1.33-1.33) in United States, Spain, Brazil, and United Kingdom, respectively, which occurred between May and early June. The corresponding threshold of vaccine ranged between 19-39%. The values of R0/ Re was toward to below 1 after August, 2022. Though the informative priors (8.5 days) was applied, the estimated infectious time before isolation ranged from 2.05 to 2.74 days (reciprocal α).

**Table 1.** Results of estimated parameters from Monkeypox outbreaks by using the Bayesian SIR model.

| Data | Total cases | The periods | Estimated by the Bayesian SIR model |  |  |  |
| --- | --- | --- | --- | --- | --- | --- |
| | | | $\beta$<br>(95% CI) | $\alpha$<br>(95% CI);<br>reciprocal $\alpha$<br>(95% CI) | $R_0/R_e$<br>(95% CI) | $V_c$ |
| The Globe | 65,215 | January 31 ~ April 30 | 0.374 (0.368-0.384) | 0.373 (0.373-0.374) | 1.001 (0.986-1.150) |  |
|  |  | May 1 ~ May 31 | 0.545 (0.511-0.563) |  | 1.459 (1.370-1.507) | 37% |
|  |  | June 1 ~ July 31 | 0.420 (0.419-0.421) |  | 1.125 (1.123-1.127) | 13% |
|  |  | August 1 ~ September 23 | 0.383 (0.383-0.384) | 2.678 (2.677-2.680) | 1.027 (1.026-1.028) | 3% |
| United States | 24,403 | May 17 ~ June 30 | 0.583 (0.581-0.589) | 0.487 (0.485-0.490) | 1.198 (1.197-1.201) | 19% |
|  |  | July 1 ~ July 30 | 0.567 (0.563-0.569) |  | 1.164 (1.149-1.169) | 17% |
|  |  | July 31 ~ August 31 | 0.499 (0.493-0.516) |  | 1.024 (1.016-1.052) | 3% |
|  |  | September 1 ~ September 23 | 0.407 (0.192-0.452) | 2.054 (2.039-2.061) | 0.837 (-0.391-0.929) | - |
| Spain | 7,083 | May 18 ~ June 16 | 0.480 (0.478-0.483) | 0.403 (0.400-0.405) | 1.191 (1.189-1.194) | 19% |
|  |  | June 17 ~ July 16 | 0.454 (0.451-0.457) |  | 1.126 (1.124-1.129) | 13% |
|  |  | July 17 ~ August 31 | 0.389 (0.386-0.391) |  | 0.963 (0.961-0.966) | - |
|  |  | September 1 ~ September 23 | 0.298 (0.278-0.316) | 2.479 (2.467-2.498) | 0.739 (0.690-0.784) | - |
| Brazil | 7,300 | June 8 ~ July 7 | 0.489 (0.487-0.492) | 0.365 (0.363-0.368) | 1.341 (1.337-1.345) | 30% |
|  |  | July 8 ~ August 6 | 0.435 (0.433-0.439) |  | 1.193 (1.189-1.197) | 19% |
|  |  | August 7 ~ September 23 | 0.362 (0.359-0.365) | 2.740 (2.717-2.758) | 0.992 (0.990-0.994) | - |
| United Kingdom | 3,585 | June 8 ~ July 7 | 0.542 (0.539-0.545) | 0.408 (0.405-0.411) | 1.330 (1.326-1.334) | 29% |
|  |  | July 8 ~ August 6 | 0.437 (0.433-0.440) |  | 1.071 (1.067-1.074) | 8% |
|  |  | August 7 ~ August 31 | 0.395 (0.392-0.398) |  | 0.969 (0.966-0.972) | - |
|  |  | September 1 ~ September 23 | 0.181 (0.115-0.280) | 2.453 (2.436-2.472) | 0.443 (0.284-0.687) | - |

$R_0$ : basic reproductive number; $R_e$ : effective reproductive number; SIR model: *Susceptible-Infected-Recovered* model.CI: credible interval

The cumulated observed cases and predicted cases by the Bayesian SIR model in different countries is shown in Figure 2. We also predicted the cumulated cases in September 30, 2022 by parameters of last period for the model validation, and we can realize the trend of Monkeypox cases.

**Figure 2.**
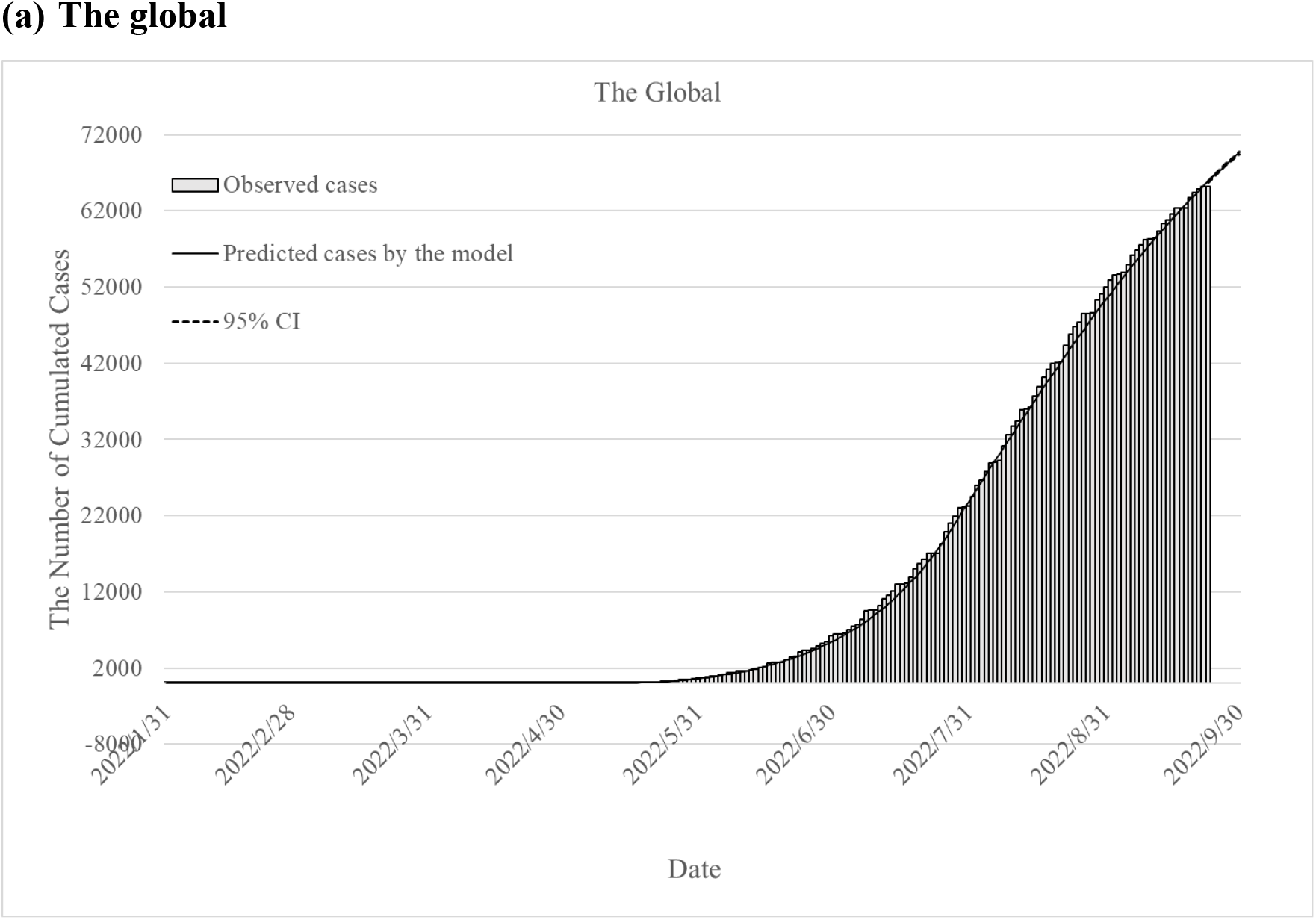
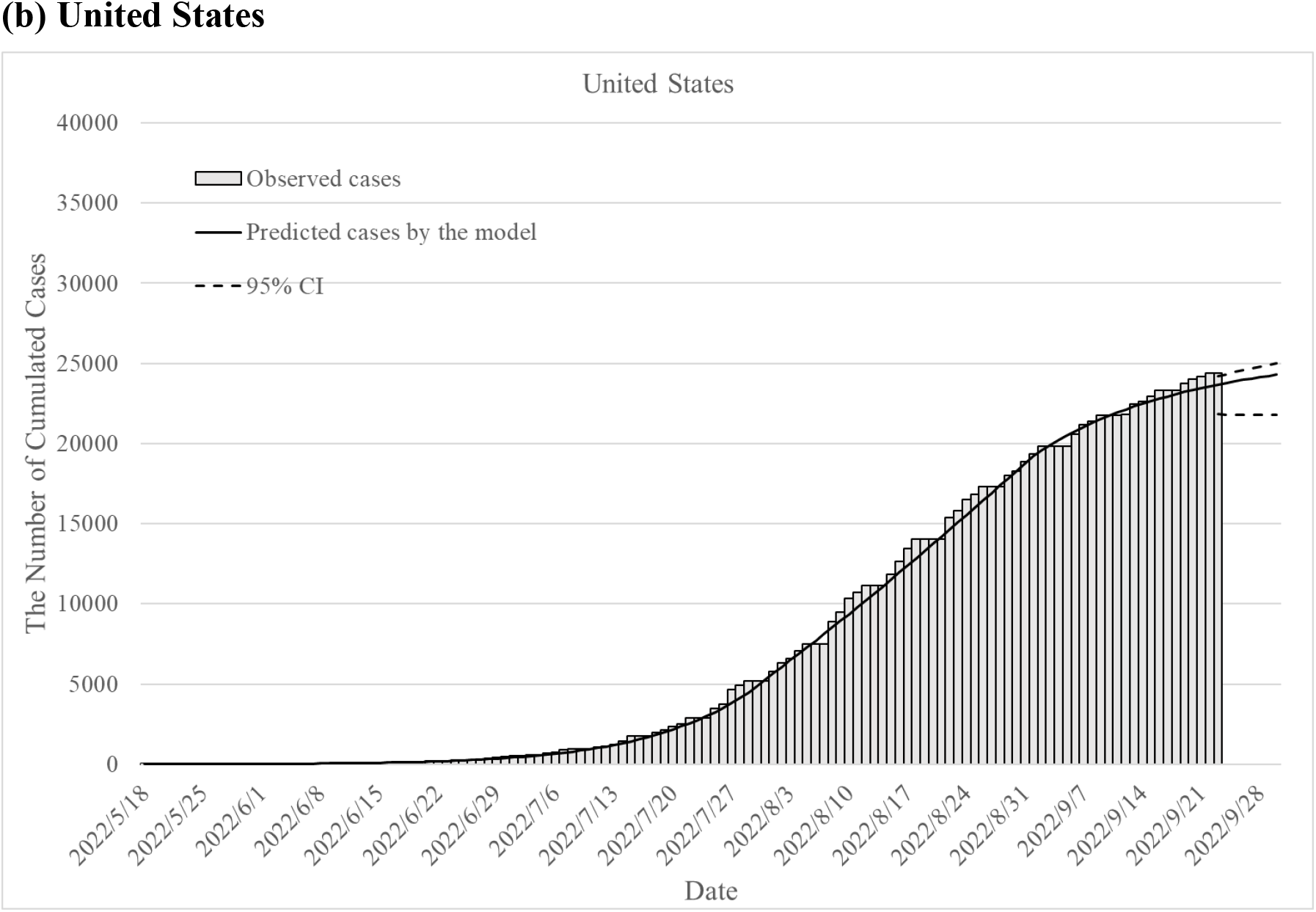
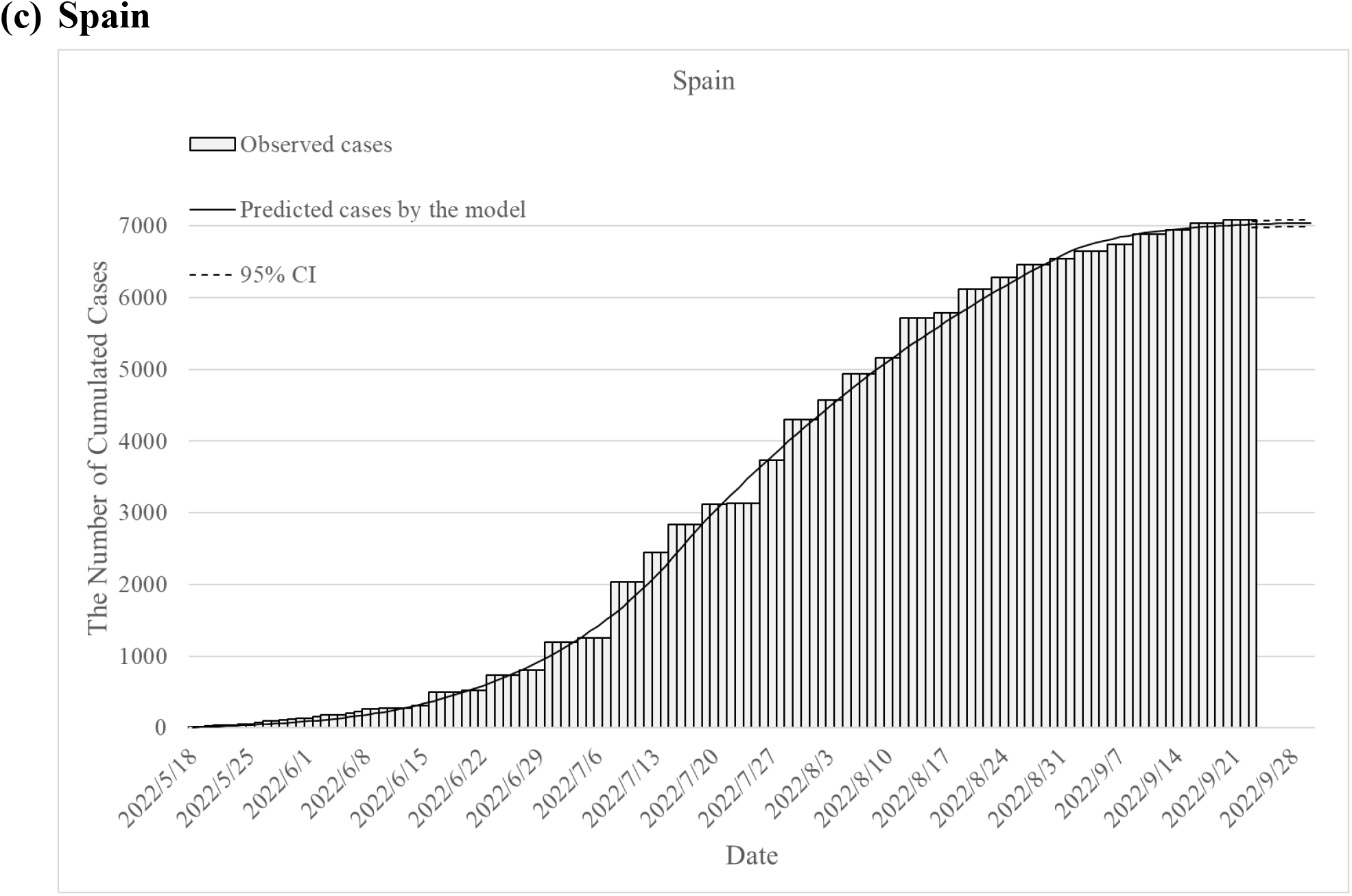
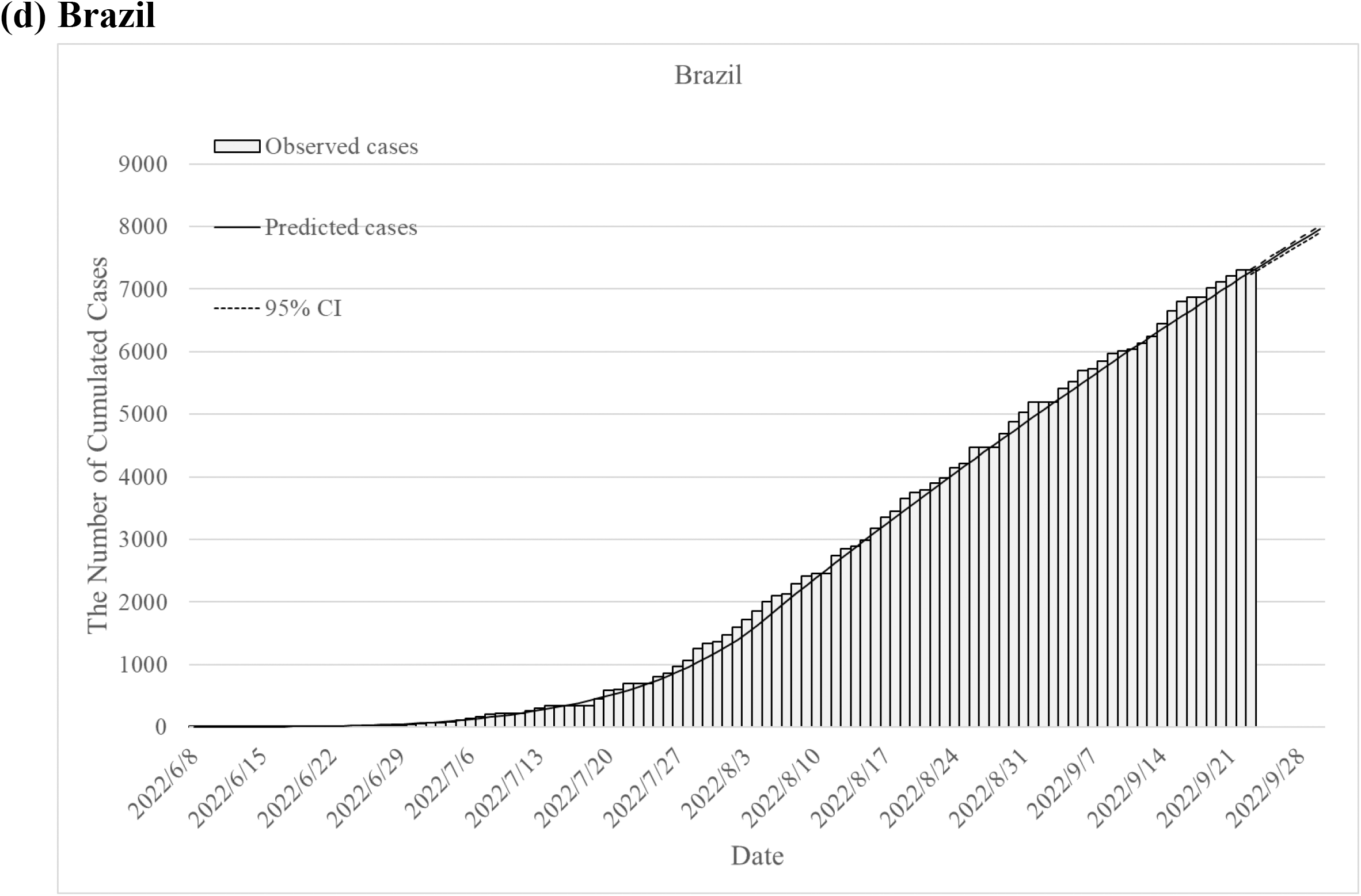
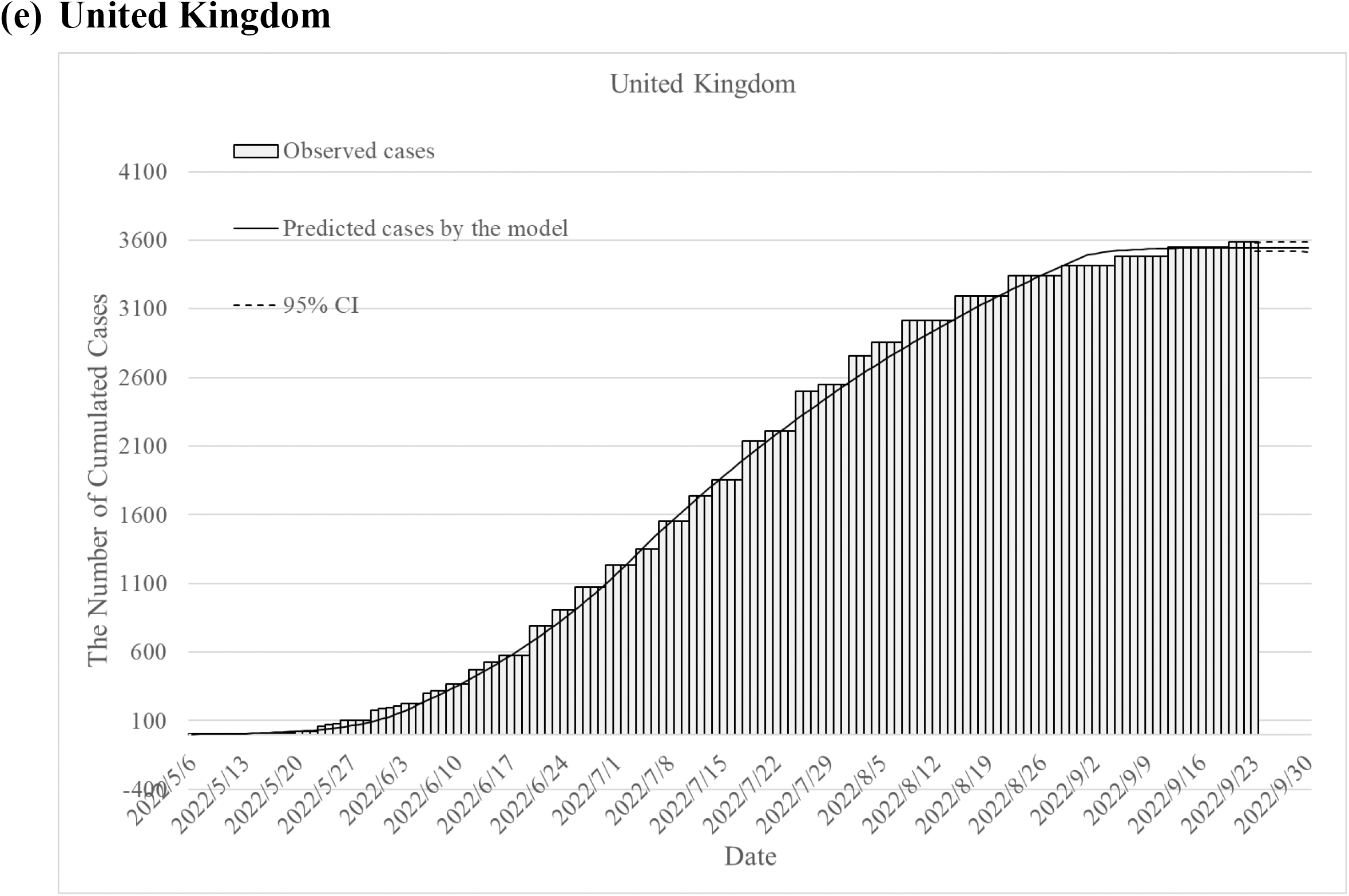
The observed cumulative Monkeypox cases and predicted cumulated Monkeypox cases on the global and in different countries.

## Discussion

The Monkeypox outbreaks seemed to control in the U.S., Spain, Brazil, and U.K., and the global because the estimated R_0_/R_e_ was gradually toward to below 1. Though it has long incubation period (up to 21 days) and long contagious period, the estimated infectious time before isolation was short.

The basic reproduction number (R_0_) or the effective reproduction number (R_e_) is defined as the expected number of secondary infections per primary infection. It often used to quantify the ability of an infectious disease to invade a population. R_0_, refers to the susceptible population without NPIs (nonpharmaceutical interventions) and vaccination, and R_e_ refers to the susceptible population with vaccination or some NPIs. R_0_/R_e_ above 1 indicates that the infectious disease is potential epidemic. R_0_/R_e_ below 1 indicates that the infectious disease is potential extinguished. The estimated R_0_/R_e_ was above one in the early stage of outbreaks, but it was below 1.5 in different countries in this study. In one previous study, 1.29 of the reproduction number was reported as of July 22, 2022. (12) It is consistent with our results.

Monkeypox and smallpox belong to the orthopoxvirus family. Both of them can be transmitted by droplet exposure, close contact with infected skin lesions or contaminated materials. The R_0_ for smallpox was estimated between 3.5 and 6.0 in the previous study. (13) However, the spreading of monkeypox is mainly through close or intimate contact with ill persons or animals. Hence, the R_0_ for monkeypox is lower than smallpox.

The spread of monkeypox among persons in this global outbreak was well controlled by early diagnosed with isolation because the estimated infectious period before cases-finding was below 3 days. Though the incubation period of monkeypox is usually from 6 to 13 days but can range from 5 to 21 days. In addition, high risk population get vaccinated to prevent from the transmission of monkeypox.

There were several limitations to estimate the R_0_ of monkeypox because a homogeneous random mixing population was assumed, the surveillance systems were the same in different countries, and the transmission of monkeypox cases was human to human outside Africa in this study. We estimated the parameters based on the Bayesian SIR model for the uncertainty to minimize these biases, and results were consistent in different countries. The estimated results were similar to the simulation data by the deterministic SIR model (supplementary).

## Conclusion

In conclusion, the global spreading of human Monkeypox outside Africa is not a threat because the R_0_/R_e_ is below one according to Bayesian reasoning epidemic model assessment. The epidemic has been therefore contained and will not result in a pandemic.

## Data Availability

The data on the Monkeypox outbreaks was retrieved from the Global health team for estimating of R0/Re. The data of the global, United States, Spain, Brazil, and United Kingdom retrieved till September 23,2022.

https://www.monkeypox.global.health/

https://www.cdc.gov/poxvirus/monkeypox/response/2022/us-map.html.

## Acknowledgements

The authors received thank the financial support from the Ministry of Science and Technology, Taiwan (MOST 111-2321-B-002-017).

## Handling editor

Chao-Chih Lai and Hsiu-Hsi Chen

## Contributors

LCL and HHC conceptualized the manuscript and LCL analyzed the data and wrote the first draft of the manuscript. LCL and CCL collected data. CYH, CCL, and HHC revised it critically for important content and validated the results. All authors agreed with the conclusions of this article. The corresponding author attests that all listed authors meet authorship criteria.

## Data availability statement

All data relevant to the study are included in the article.

## Ethics approval

Ethical approval was not required because this study does not involve personal information, all data are compiled from verified sources, including reports from governments and public health organizations and news media reporting of health official statements.

## Patient consent for publication

Not applicable

