## Supplemental for "Estimating the Global Spread of Epidemic Human Monkeypox with Bayesian Directed Acyclic Graphic Model"

**Supplementary:**

The data of monkeypox outbreaks are listed in Table A1. The results of predicted monkeypox cases by Bayesian SIR model are also listed in Table A1.

**The deterministic SIR model**

A deterministic Susceptible-Infected-Recovered (SIR) model was also used for the Monkeypox outbreaks.

Nonlinear ordinary differential equations of the COVID-19 model were used to describe the dynamic changes of each state as follows.

$$\frac{dS(t)}{dt} = -\beta \frac{S(t)}{N} I(t) \quad (1)$$

$$\frac{dI(t)}{dt} = \beta \frac{S(t)}{N} I(t) - \alpha I(t) \quad (2)$$

$$\frac{dR(t)}{dt} = \alpha I(t) \quad (3)$$

Where β denotes the transmission coefficient, overall transmissibility with contact rate; α denotes reciprocal of the infectious period (8.5 days) and it was set to be 0.1176. The formula of deriving the basic reproductive number ($R_0$) is $\frac{\beta}{\alpha}$ . The β and $R_0$ were obtained after fitting the observed cases with monkeypox by the simulation of this model by deterministic SEIR model. The assumptions of our deterministic SEIR model are as follows. (1) Homogeneous random mixing population; (2) fixed infectious period. The simulation of the deterministic SEIR model was performed by using the software of MATLAB version 9.30.713579 (MathWorks, USA).

The results of estimated parameters for monkeypox in the global, United States, Spain, Brazil, and United Kingdom are listed in Table A2. The observed cumulative monkeypox cases and simulated cumulated Monkeypox cases on the global, United States, Spain, Brazil, and United Kingdom are shown in Figure A1. The results of simulated monkeypox cases by the deterministic SIR model are listed in Table A3.

**Table A1 The observed cumulated monkeypox cases and predicted cumulated cases by the Bayesian SIR model**

**(a) The global monkeypox cases**

| Date | Observed cumulated cases | Predicted cumulated cases | 95% CI | | Date | Observed cumulated cases | Predicted cumulated cases | 95% CI | |
|---|---|---|---|---|---|---|---|---|---|
| 2022/1/31 | 3 | 1.0 | - | - | 2022/5/8 | 37 | 45.2 | 32.6 | 76.4 |
| 2022/2/16 | 3 | 7.0 | 6.7 | 7.7 | 2022/5/9 | 37 | 47.3 | 34.0 | 80.3 |
| 2022/2/17 | 6 | 7.4 | 7.0 | 8.2 | 2022/5/10 | 37 | 49.9 | 35.8 | 84.8 |
| 2022/2/27 | 6 | 11.2 | 10.3 | 13.1 | 2022/5/11 | 37 | 52.9 | 37.9 | 89.9 |
| 2022/2/28 | 7 | 11.6 | 10.6 | 13.6 | 2022/5/12 | 38 | 56.5 | 40.5 | 95.8 |
| 2022/3/3 | 7 | 12.7 | 11.5 | 15.2 | 2022/5/13 | 39 | 60.7 | 43.5 | 102.6 |
| 2022/3/4 | 9 | 13.1 | 11.9 | 15.7 | 2022/5/14 | 39 | 65.7 | 47.3 | 110.3 |
| 2022/3/30 | 9 | 23.4 | 19.4 | 32.2 | 2022/5/15 | 43 | 71.6 | 51.8 | 119.2 |
| 2022/3/31 | 15 | 23.8 | 19.7 | 32.9 | 2022/5/16 | 43 | 78.6 | 57.3 | 129.5 |
| 2022/4/9 | 15 | 27.5 | 22.1 | 39.9 | 2022/5/17 | 46 | 86.8 | 63.9 | 141.2 |
| 2022/4/10 | 19 | 27.9 | 22.3 | 40.7 | 2022/5/18 | 67 | 96.6 | 71.8 | 154.7 |
| 2022/4/11 | 19 | 28.3 | 22.6 | 41.6 | 2022/5/19 | 86 | 108.1 | 81.4 | 170.1 |
| 2022/4/12 | 21 | 28.7 | 22.8 | 42.4 | 2022/5/20 | 131 | 121.8 | 93.1 | 187.9 |
| 2022/4/13 | 21 | 29.2 | 23.1 | 43.2 | 2022/5/21 | 147 | 138.0 | 107.1 | 208.3 |
| 2022/4/14 | 21 | 29.6 | 23.3 | 44.1 | 2022/5/22 | 147 | 157.2 | 124.0 | 231.7 |
| 2022/4/15 | 21 | 30.0 | 23.6 | 45.0 | 2022/5/23 | 220 | 179.9 | 144.5 | 258.6 |
| 2022/4/16 | 21 | 30.4 | 23.8 | 45.8 | 2022/5/24 | 256 | 206.9 | 169.2 | 289.4 |
| 2022/4/17 | 21 | 30.9 | 24.1 | 46.7 | 2022/5/25 | 300 | 238.8 | 199.1 | 324.9 |
| 2022/4/18 | 21 | 31.3 | 24.3 | 47.6 | 2022/5/26 | 388 | 276.7 | 235.1 | 365.5 |
| 2022/4/19 | 21 | 31.7 | 24.6 | 48.5 | 2022/5/27 | 440 | 321.6 | 278.6 | 412.2 |
| 2022/4/20 | 21 | 32.1 | 24.8 | 49.4 | 2022/5/28 | 457 | 374.9 | 331.0 | 465.8 |
| 2022/4/21 | 21 | 32.6 | 25.1 | 50.4 | 2022/5/29 | 477 | 438.1 | 395.0 | 527.8 |
| 2022/4/22 | 21 | 33.0 | 25.3 | 51.3 | 2022/5/30 | 600 | 513.1 | 471.8 | 598.2 |
| 2022/4/23 | 21 | 33.4 | 25.6 | 52.2 | 2022/5/31 | 665 | 602.2 | 564.2 | 678.8 |
| 2022/4/24 | 21 | 33.8 | 25.8 | 53.2 | 2022/6/1 | 749 | 678.6 | 641.7 | 751.9 |
| 2022/4/25 | 21 | 34.3 | 26.0 | 54.1 | 2022/6/2 | 874 | 758.7 | 722.9 | 828.5 |
| 2022/4/26 | 21 | 34.7 | 26.3 | 55.1 | 2022/6/3 | 968 | 842.5 | 808.1 | 908.9 |
| 2022/4/27 | 21 | 35.1 | 26.5 | 56.1 | 2022/6/4 | 976 | 930.3 | 897.9 | 993.8 |
| 2022/4/28 | 21 | 35.6 | 26.7 | 57.1 | 2022/6/5 | 1083 | 1022.2 | 990.9 | 1082.0 |
| 2022/4/29 | 21 | 36.0 | 27.0 | 58.1 | 2022/6/6 | 1198 | 1118.5 | 1089.3 | 1175.3 |
| 2022/4/30 | 26 | 36.5 | 27.2 | 59.1 | 2022/6/7 | 1348 | 1219.3 | 1190.6 | 1271.5 |
| 2022/5/1 | 26 | 36.9 | 27.4 | 60.2 | 2022/6/8 | 1445 | 1324.8 | 1299.9 | 1375.4 |
| 2022/5/2 | 26 | 37.6 | 27.8 | 61.6 | 2022/6/9 | 1591 | 1435.3 | 1411.8 | 1481.5 |
| 2022/5/3 | 26 | 38.4 | 28.3 | 63.3 | 2022/6/10 | 1634 | 1551.0 | 1528.1 | 1592.0 |
| 2022/5/4 | 26 | 39.3 | 28.9 | 65.3 | 2022/6/11 | 1643 | 1672.1 | 1650.5 | 1708.5 |
| 2022/5/5 | 26 | 40.5 | 29.5 | 67.5 | 2022/6/12 | 1757 | 1798.9 | 1778.7 | 1830.7 |
| 2022/5/6 | 27 | 41.8 | 30.4 | 70.1 | 2022/6/13 | 1817 | 1931.6 | 1912.9 | 1959.2 |
| 2022/5/7 | 27 | 43.3 | 31.4 | 73.1 | 2022/6/14 | 2061 | 2070.5 | 2053.9 | 2094.1 |

CI: credible interval

| Date | Observed cumulated cases | Predicted cumulated cases | 95% CI | | Date | Observed cumulated cases | Predicted cumulated cases | 95% CI | |
|---|---|---|---|---|---|---|---|---|---|
| 2022/6/15 | 2208 | 2215.8 | 2200.9 | 2235.1 | 2022/7/23 | 17016 | 16773.5 | 16664.6 | 16830.9 |
| 2022/6/16 | 2584 | 2368.0 | 2354.1 | 2383.6 | 2022/7/24 | 17050 | 17550.3 | 17447.8 | 17606.1 |
| 2022/6/17 | 2778 | 2527.2 | 2515.5 | 2540.4 | 2022/7/25 | 18336 | 18357.5 | 18261.8 | 18411.5 |
| 2022/6/18 | 2783 | 2693.7 | 2682.6 | 2705.0 | 2022/7/26 | 19872 | 19196.0 | 19109.1 | 19248.5 |
| 2022/6/19 | 2790 | 2868.1 | 2856.2 | 2879.1 | 2022/7/27 | 21025 | 20066.7 | 19987.4 | 20116.0 |
| 2022/6/20 | 3084 | 3050.4 | 3036.4 | 3063.2 | 2022/7/28 | 21867 | 20970.4 | 20900.0 | 21015.6 |
| 2022/6/21 | 3366 | 3241.2 | 3223.6 | 3256.3 | 2022/7/29 | 23087 | 21908.0 | 21849.5 | 21949.4 |
| 2022/6/22 | 3540 | 3440.7 | 3419.1 | 3458.6 | 2022/7/30 | 23172 | 22880.4 | 22833.5 | 22917.4 |
| 2022/6/23 | 4098 | 3649.5 | 3623.8 | 3670.2 | 2022/7/31 | 23303 | 23784.5 | 23746.5 | 23820.9 |
| 2022/6/24 | 4303 | 3867.8 | 3836.1 | 3890.2 | 2022/8/1 | 24521 | 24688.6 | 24655.1 | 24720.5 |
| 2022/6/25 | 4315 | 4096.0 | 4058.6 | 4120.3 | 2022/8/2 | 25963 | 25592.4 | 25563.8 | 25623.2 |
| 2022/6/26 | 4511 | 4334.7 | 4292.7 | 4362.1 | 2022/8/3 | 26616 | 26495.5 | 26467.8 | 26522.7 |
| 2022/6/27 | 4898 | 4584.3 | 4535.1 | 4612.2 | 2022/8/4 | 27740 | 27397.8 | 27370.9 | 27424.3 |
| 2022/6/28 | 5246 | 4845.2 | 4792.0 | 4876.7 | 2022/8/5 | 28907 | 28298.8 | 28270.2 | 28324.7 |
| 2022/6/29 | 5399 | 5117.9 | 5058.8 | 5151.2 | 2022/8/6 | 29021 | 29198.3 | 29167.8 | 29225.8 |
| 2022/6/30 | 6206 | 5402.9 | 5338.6 | 5438.7 | 2022/8/7 | 29193 | 30096.0 | 30065.9 | 30128.9 |
| 2022/7/1 | 6475 | 5700.7 | 5631.4 | 5739.0 | 2022/8/8 | 31178 | 30991.5 | 30956.0 | 31025.3 |
| 2022/7/2 | 6519 | 6011.9 | 5939.3 | 6054.4 | 2022/8/9 | 32613 | 31884.5 | 31848.1 | 31923.1 |
| 2022/7/3 | 6559 | 6337.0 | 6258.9 | 6381.3 | 2022/8/10 | 33766 | 32774.8 | 32735.7 | 32816.2 |
| 2022/7/4 | 7053 | 6676.6 | 6593.3 | 6723.0 | 2022/8/11 | 34451 | 33662.1 | 33622.9 | 33708.3 |
| 2022/7/5 | 7500 | 7031.2 | 6942.8 | 7079.2 | 2022/8/12 | 35863 | 34546.0 | 34504.9 | 34595.4 |
| 2022/7/6 | 7732 | 7401.6 | 7307.9 | 7451.2 | 2022/8/13 | 36004 | 35426.3 | 35380.6 | 35475.3 |
| 2022/7/7 | 8426 | 7788.2 | 7690.0 | 7839.7 | 2022/8/14 | 36172 | 36302.7 | 36256.9 | 36355.9 |
| 2022/7/8 | 9486 | 8191.8 | 8097.0 | 8252.5 | 2022/8/15 | 37646 | 37175.0 | 37127.1 | 37229.8 |
| 2022/7/9 | 9608 | 8613.0 | 8514.7 | 8675.6 | 2022/8/16 | 38965 | 38042.8 | 37994.3 | 38100.1 |
| 2022/7/10 | 9635 | 9052.5 | 8944.2 | 9110.2 | 2022/8/17 | 40147 | 38906.0 | 38859.8 | 38968.8 |
| 2022/7/11 | 10226 | 9511.0 | 9405.5 | 9576.2 | 2022/8/18 | 41192 | 39764.2 | 39717.5 | 39828.6 |
| 2022/7/12 | 11098 | 9989.2 | 9880.2 | 10055.3 | 2022/8/19 | 41964 | 40617.2 | 40570.3 | 40683.2 |
| 2022/7/13 | 11516 | 10487.8 | 10372.8 | 10551.0 | 2022/8/20 | 42130 | 41464.8 | 41417.4 | 41531.5 |
| 2022/7/14 | 12127 | 11007.7 | 10890.6 | 11071.4 | 2022/8/21 | 42238 | 42306.7 | 42259.1 | 42374.0 |
| 2022/7/15 | 13015 | 11549.6 | 11431.3 | 11614.5 | 2022/8/22 | 44301 | 43142.7 | 43091.1 | 43206.7 |
| 2022/7/16 | 13031 | 12114.2 | 11996.4 | 12180.8 | 2022/8/23 | 45752 | 43972.6 | 43920.8 | 44036.2 |
| 2022/7/17 | 13069 | 12702.4 | 12584.1 | 12768.7 | 2022/8/24 | 46801 | 44796.1 | 44744.9 | 44859.7 |
| 2022/7/18 | 13886 | 13315.1 | 13196.6 | 13381.0 | 2022/8/25 | 47438 | 45613.1 | 45562.9 | 45676.6 |
| 2022/7/19 | 15081 | 13952.9 | 13834.9 | 14018.1 | 2022/8/26 | 48504 | 46423.4 | 46377.8 | 46489.9 |
| 2022/7/20 | 15655 | 14616.8 | 14499.7 | 14680.4 | 2022/8/27 | 48563 | 47226.8 | 47176.6 | 47286.8 |
| 2022/7/21 | 16293 | 15307.6 | 15194.3 | 15371.5 | 2022/8/28 | 48616 | 48023.1 | 47979.4 | 48087.4 |
| 2022/7/22 | 17013 | 16026.2 | 15912.4 | 16084.7 | 2022/8/29 | 50343 | 48812.0 | 48768.3 | 48873.4 |

CI: credible interval

| Date | Observed cumulated cases | Predicted cumulated cases | 95% CI | |
|---|---|---|---|---|
| 2022/8/30 | 51110 | 49593.6 | 49547.9 | 49649.9 |
| 2022/8/31 | 51982 | 50367.5 | 50322.0 | 50420.1 |
| 2022/9/1 | 52941 | 51133.8 | 51090.5 | 51185.1 |
| 2022/9/2 | 53629 | 51892.1 | 51849.2 | 51939.6 |
| 2022/9/3 | 53678 | 52642.4 | 52602.2 | 52688.4 |
| 2022/9/4 | 53936 | 53384.6 | 53345.8 | 53427.7 |
| 2022/9/5 | 54939 | 54118.5 | 54077.4 | 54155.9 |
| 2022/9/6 | 56138 | 54844.1 | 54806.8 | 54881.3 |
| 2022/9/7 | 56852 | 55561.2 | 55524.3 | 55595.7 |
| 2022/9/8 | 57490 | 56269.7 | 56235.5 | 56304.6 |
| 2022/9/9 | 58211 | 56969.6 | 56935.9 | 57002.6 |
| 2022/9/10 | 58322 | 57660.9 | 57628.6 | 57693.7 |
| 2022/9/11 | 58392 | 58343.3 | 58309.4 | 58374.5 |
| 2022/9/12 | 59351 | 59017.0 | 58983.0 | 59049.9 |
| 2022/9/13 | 60337 | 59681.7 | 59646.9 | 59716.3 |
| 2022/9/14 | 60784 | 60337.5 | 60300.3 | 60375.1 |
| 2022/9/15 | 61581 | 60984.4 | 60944.4 | 61025.7 |
| 2022/9/16 | 62324 | 61622.2 | 61578.9 | 61667.5 |
| 2022/9/17 | 62399 | 62251.1 | 62202.8 | 62300.0 |
| 2022/9/18 | 62403 | 62870.9 | 62815.3 | 62920.6 |
| 2022/9/19 | 63758 | 63481.7 | 63420.2 | 63534.9 |
| 2022/9/20 | 64418 | 64083.4 | 64021.6 | 64145.4 |
| 2022/9/21 | 64904 | 64676.1 | 64605.5 | 64738.6 |
| 2022/9/22 | 65215 | 65259.8 | 65182.8 | 65326.4 |
| 2022/9/23 | 65215 | 65834.4 | 65753.8 | 65907.8 |
| 2022/9/24 | | 66400.0 | 66312.6 | 66477.1 |
| 2022/9/25 | | 66956.7 | 66861.8 | 67037.1 |
| 2022/9/26 | | 67504.4 | 67402.0 | 67588.7 |
| 2022/9/27 | | 68043.2 | 67933.1 | 68131.6 |
| 2022/9/28 | | 68573.2 | 68456.1 | 68666.4 |
| 2022/9/29 | | 69094.3 | 68971.4 | 69193.5 |
| 2022/9/30 | | 69606.6 | 69476.1 | 69710.3 |

CI: credible interval

### (b) United States

| Date | Observed cumulated cases | Predicted cumulated cases | 95% CI | | Date | Observed cumulated cases | Predicted cumulated cases | 95% CI | |
|---|---|---|---|---|---|---|---|---|---|
| 2022/5/18 | 1 | 1.0 | | | 2022/6/25 | 243 | 232.0 | 225.7 | 247.3 |
| 2022/5/19 | 2 | 1.6 | 1.6 | 1.6 | 2022/6/26 | 252 | 256.0 | 248.9 | 273.4 |
| 2022/5/20 | 2 | 2.3 | 2.3 | 2.3 | 2022/6/27 | 315 | 282.5 | 274.4 | 302.1 |
| 2022/5/21 | 2 | 3.0 | 3.0 | 3.1 | 2022/6/28 | 359 | 311.6 | 302.4 | 333.9 |
| 2022/5/22 | 2 | 3.8 | 3.8 | 3.9 | 2022/6/29 | 409 | 343.7 | 333.3 | 368.9 |
| 2022/5/23 | 2 | 4.7 | 4.7 | 4.8 | 2022/6/30 | 470 | 379.0 | 367.3 | 407.6 |
| 2022/5/24 | 4 | 5.7 | 5.7 | 5.8 | 2022/7/1 | 543 | 417.9 | 404.6 | 450.2 |
| 2022/5/25 | 6 | 6.8 | 6.8 | 6.9 | 2022/7/2 | 547 | 459.1 | 444.4 | 494.7 |
| 2022/5/26 | 11 | 8.0 | 8.0 | 8.2 | 2022/7/3 | 553 | 503.7 | 487.5 | 542.4 |
| 2022/5/27 | 15 | 9.4 | 9.3 | 9.5 | 2022/7/4 | 563 | 552.0 | 534.4 | 593.8 |
| 2022/5/28 | 16 | 10.8 | 10.8 | 11.0 | 2022/7/5 | 709 | 604.3 | 585.6 | 649.4 |
| 2022/5/29 | 17 | 12.4 | 12.3 | 12.7 | 2022/7/6 | 749 | 660.9 | 640.4 | 708.5 |
| 2022/5/30 | 17 | 14.2 | 14.1 | 14.5 | 2022/7/7 | 883 | 722.1 | 700.3 | 772.5 |
| 2022/5/31 | 21 | 16.2 | 16.0 | 16.5 | 2022/7/8 | 950 | 788.4 | 765.2 | 841.2 |
| 2022/6/1 | 22 | 18.3 | 18.1 | 18.8 | 2022/7/9 | 960 | 860.2 | 835.7 | 915.2 |
| 2022/6/2 | 26 | 20.7 | 20.5 | 21.2 | 2022/7/10 | 964 | 937.9 | 912.2 | 994.9 |
| 2022/6/3 | 31 | 23.3 | 23.0 | 23.9 | 2022/7/11 | 1058 | 1021.9 | 995.0 | 1080.3 |
| 2022/6/4 | 32 | 26.2 | 25.9 | 26.9 | 2022/7/12 | 1122 | 1112.9 | 1085.2 | 1172.7 |
| 2022/6/5 | 32 | 29.3 | 29.0 | 30.2 | 2022/7/13 | 1245 | 1211.3 | 1182.7 | 1271.7 |
| 2022/6/6 | 34 | 32.8 | 32.4 | 33.9 | 2022/7/14 | 1426 | 1317.8 | 1288.4 | 1378.1 |
| 2022/6/7 | 40 | 36.6 | 36.2 | 37.9 | 2022/7/15 | 1767 | 1433.1 | 1403.1 | 1492.8 |
| 2022/6/8 | 45 | 40.9 | 40.3 | 42.3 | 2022/7/16 | 1767 | 1557.7 | 1527.4 | 1615.8 |
| 2022/6/9 | 50 | 45.5 | 44.9 | 47.2 | 2022/7/17 | 1767 | 1692.6 | 1662.2 | 1748.3 |
| 2022/6/10 | 53 | 50.7 | 49.9 | 52.6 | 2022/7/18 | 1995 | 1838.6 | 1808.4 | 1890.8 |
| 2022/6/11 | 55 | 56.3 | 55.4 | 58.6 | 2022/7/19 | 2114 | 1996.4 | 1968.9 | 2046.0 |
| 2022/6/12 | 57 | 62.5 | 61.5 | 65.2 | 2022/7/20 | 2323 | 2167.2 | 2139.6 | 2209.4 |
| 2022/6/13 | 75 | 69.4 | 68.1 | 72.4 | 2022/7/21 | 2503 | 2351.8 | 2326.8 | 2387.1 |
| 2022/6/14 | 83 | 76.9 | 75.5 | 80.4 | 2022/7/22 | 2875 | 2551.5 | 2528.9 | 2578.6 |
| 2022/6/15 | 96 | 85.2 | 83.6 | 89.3 | 2022/7/23 | 2875 | 2767.4 | 2748.0 | 2786.5 |
| 2022/6/16 | 117 | 94.4 | 92.5 | 99.0 | 2022/7/24 | 2875 | 3000.8 | 2980.0 | 3019.1 |
| 2022/6/17 | 128 | 104.4 | 102.3 | 109.8 | 2022/7/25 | 3457 | 3253.0 | 3217.8 | 3275.9 |
| 2022/6/18 | 132 | 115.5 | 113.0 | 121.6 | 2022/7/26 | 3768 | 3525.7 | 3472.5 | 3555.0 |
| 2022/6/19 | 133 | 127.8 | 124.9 | 134.7 | 2022/7/27 | 4630 | 3820.3 | 3744.1 | 3855.4 |
| 2022/6/20 | 137 | 141.2 | 138.0 | 149.2 | 2022/7/28 | 4895 | 4138.7 | 4034.9 | 4180.3 |
| 2022/6/21 | 175 | 156.1 | 152.3 | 165.2 | 2022/7/29 | 5175 | 4482.5 | 4346.4 | 4531.8 |
| 2022/6/22 | 196 | 172.4 | 168.1 | 182.8 | 2022/7/30 | 5175 | 4853.8 | 4681.1 | 4913.4 |
| 2022/6/23 | 218 | 190.4 | 185.5 | 202.2 | 2022/7/31 | 5175 | 5254.7 | 5041.7 | 5327.3 |
| 2022/6/24 | 237 | 210.2 | 204.7 | 223.6 | 2022/8/1 | 5792 | 5687.5 | 5427.4 | 5774.6 |

CI: credible interval

| Date | Observed cumulated cases | Predicted cumulated cases | 95% CI | | Date | Observed cumulated cases | Predicted cumulated cases | 95% CI | |
|---|---|---|---|---|---|---|---|---|---|
| 2022/8/2 | 6308 | 6084.3 | 5796.5 | 6180.3 | 2022/9/9 | 21761 | 21519.0 | 21387.3 | 21871.9 |
| 2022/8/3 | 6599 | 6484.3 | 6171.6 | 6585.8 | 2022/9/10 | 21761 | 21732.8 | 21646.8 | 21967.1 |
| 2022/8/4 | 7084 | 6887.5 | 6556.6 | 6995.1 | 2022/9/11 | 21761 | 21934.8 | 21770.5 | 22106.9 |
| 2022/8/5 | 7490 | 7293.8 | 6951.5 | 7407.6 | 2022/9/12 | 21834 | 22126.1 | 21781.5 | 22247.1 |
| 2022/8/6 | 7491 | 7703.3 | 7352.4 | 7818.8 | 2022/9/13 | 22480 | 22307.2 | 21803.6 | 22438.5 |
| 2022/8/7 | 7491 | 8115.7 | 7763.5 | 8233.0 | 2022/9/14 | 22619 | 22478.7 | 21807.6 | 22647.5 |
| 2022/8/8 | 8902 | 8531.2 | 8182.7 | 8648.3 | 2022/9/15 | 22958 | 22641.4 | 21813.0 | 22853.9 |
| 2022/8/9 | 9461 | 8949.6 | 8611.6 | 9066.0 | 2022/9/16 | 23339 | 22795.6 | 21820.2 | 23052.5 |
| 2022/8/10 | 10361 | 9370.9 | 9047.6 | 9483.5 | 2022/9/17 | 23339 | 22941.9 | 21819.0 | 23235.6 |
| 2022/8/11 | 10727 | 9795.1 | 9496.3 | 9905.8 | 2022/9/18 | 23339 | 23080.8 | 21824.4 | 23419.3 |
| 2022/8/12 | 11131 | 10222.0 | 9951.6 | 10327.6 | 2022/9/19 | 23730 | 23212.6 | 21820.6 | 23585.8 |
| 2022/8/13 | 11131 | 10651.7 | 10413.1 | 10747.0 | 2022/9/20 | 24041 | 23337.7 | 21818.7 | 23747.6 |
| 2022/8/14 | 11131 | 11084.0 | 10881.8 | 11168.8 | 2022/9/21 | 24198 | 23456.5 | 21826.9 | 23911.8 |
| 2022/8/15 | 11844 | 11518.9 | 11359.2 | 11593.8 | 2022/9/22 | 24403 | 23569.4 | 21827.3 | 24061.7 |
| 2022/8/16 | 12636 | 11956.3 | 11837.8 | 12018.5 | 2022/9/23 | 24403 | 23676.6 | 21829.3 | 24208.1 |
| 2022/8/17 | 13450 | 12396.3 | 12314.2 | 12445.6 | 2022/9/9 | 21761 | 21519.0 | 21387.3 | 21871.9 |
| 2022/8/18 | 14049 | 12838.6 | 12775.6 | 12896.5 | 2022/9/10 | 21761 | 21732.8 | 21646.8 | 21967.1 |
| 2022/8/19 | 14049 | 13283.2 | 13219.4 | 13402.9 | 2022/9/11 | 21761 | 21934.8 | 21770.5 | 22106.9 |
| 2022/8/20 | 14049 | 13730.1 | 13658.4 | 13933.0 | 2022/9/12 | 21834 | 22126.1 | 21781.5 | 22247.1 |
| 2022/8/21 | 14049 | 14179.2 | 14085.8 | 14471.7 | 2022/9/13 | 22480 | 22307.2 | 21803.6 | 22438.5 |
| 2022/8/22 | 15357 | 14630.5 | 14516.5 | 15020.6 | 2022/9/14 | 22619 | 22478.7 | 21807.6 | 22647.5 |
| 2022/8/23 | 15832 | 15083.7 | 14943.5 | 15577.3 | 2022/9/15 | 22958 | 22641.4 | 21813.0 | 22853.9 |
| 2022/8/24 | 16514 | 15538.9 | 15373.9 | 16146.8 | 2022/9/16 | 23339 | 22795.6 | 21820.2 | 23052.5 |
| 2022/8/25 | 16837 | 15996.0 | 15808.0 | 16730.9 | 2022/9/17 | 23339 | 22941.9 | 21819.0 | 23235.6 |
| 2022/8/26 | 17336 | 16454.9 | 16231.9 | 17312.6 | 2022/9/18 | 23339 | 23080.8 | 21824.4 | 23419.3 |
| 2022/8/27 | 17336 | 16915.4 | 16659.7 | 17910.6 | 2022/9/19 | 23730 | 23212.6 | 21820.6 | 23585.8 |
| 2022/8/28 | 17336 | 17377.7 | 17087.2 | 18519.3 | 2022/9/20 | 24041 | 23337.7 | 21818.7 | 23747.6 |
| 2022/8/29 | 17986 | 17841.4 | 17511.9 | 19133.9 | 2022/9/21 | 24198 | 23456.5 | 21826.9 | 23911.8 |
| 2022/8/30 | 18298 | 18306.6 | 17940.3 | 19762.6 | 2022/9/22 | 24403 | 23569.4 | 21827.3 | 24061.7 |
| 2022/8/31 | 18879 | 18773.1 | 18361.8 | 20394.9 | 2022/9/23 | 24403 | 23676.6 | 21829.3 | 24208.1 |
| 2022/9/1 | 19355 | 19241.0 | 18791.5 | 21046.3 | 2022/9/24 | | 23778.4 | 21813.6 | 24329.9 |
| 2022/9/2 | 19852 | 19597.6 | 19172.1 | 21254.8 | 2022/9/25 | | 23875.1 | 21813.1 | 24459.4 |
| 2022/9/3 | 19852 | 19928.9 | 19540.7 | 21409.1 | 2022/9/26 | | 23967.0 | 21813.7 | 24584.5 |
| 2022/9/4 | 19852 | 20237.8 | 19888.4 | 21516.7 | 2022/9/27 | | 24054.4 | 21802.1 | 24690.6 |
| 2022/9/5 | 19852 | 20526.8 | 20219.1 | 21596.5 | 2022/9/28 | | 24137.4 | 21802.2 | 24803.2 |
| 2022/9/6 | 20608 | 20797.9 | 20536.7 | 21660.8 | 2022/9/29 | | 24216.2 | 21800.9 | 24909.3 |
| 2022/9/7 | 21148 | 21052.7 | 20828.6 | 21714.2 | 2022/9/30 | | 24291.2 | 21802.2 | 25012.1 |
| 2022/9/8 | 21374 | 21292.6 | 21113.3 | 21781.1 | | | | | |

CI: credible interval

### (c) Spain

| Date | Observed cumulated cases | Predicted cumulated cases | 95% CI | | Date | Observed cumulated cases | Predicted cumulated cases | 95% CI | |
|---|---|---|---|---|---|---|---|---|---|
| 2022/5/18 | 7 | 7.0 | | | 2022/6/25 | 736 | 718.7 | 708.3 | 727.9 |
| 2022/5/19 | 7 | 10.5 | 10.4758 | 10.5114 | 2022/6/26 | 736 | 767.9 | 757 | 777.6 |
| 2022/5/20 | 30 | 14.3 | 14.2303 | 14.3042 | 2022/6/27 | 800 | 819.7 | 808.6 | 830.1 |
| 2022/5/21 | 40 | 18.3 | 18.2872 | 18.4028 | 2022/6/28 | 800 | 874.2 | 862.8 | 884.9 |
| 2022/5/22 | 40 | 22.8 | 22.6646 | 22.8257 | 2022/6/29 | 800 | 931.5 | 919.7 | 942.6 |
| 2022/5/23 | 41 | 27.5 | 27.3974 | 27.6092 | 2022/6/30 | 1196 | 991.7 | 979.6 | 1003.2 |
| 2022/5/24 | 45 | 32.6 | 32.506 | 32.7768 | 2022/7/1 | 1196 | 1055.1 | 1043 | 1067.4 |
| 2022/5/25 | 53 | 38.2 | 38.022 | 38.3584 | 2022/7/2 | 1196 | 1121.8 | 1109.4 | 1134.2 |
| 2022/5/26 | 78 | 44.2 | 43.9787 | 44.3912 | 2022/7/3 | 1196 | 1191.9 | 1179.5 | 1204.7 |
| 2022/5/27 | 99 | 50.7 | 50.3905 | 50.8949 | 2022/7/4 | 1258 | 1265.6 | 1253.5 | 1279 |
| 2022/5/28 | 99 | 57.7 | 57.3301 | 57.9349 | 2022/7/5 | 1258 | 1343.2 | 1330.6 | 1356.4 |
| 2022/5/29 | 108 | 65.2 | 64.8154 | 65.5441 | 2022/7/6 | 1258 | 1424.7 | 1411.9 | 1437.7 |
| 2022/5/30 | 116 | 73.4 | 72.9079 | 73.7746 | 2022/7/7 | 1258 | 1510.5 | 1497.9 | 1523.8 |
| 2022/5/31 | 136 | 82.2 | 81.6215 | 82.6531 | 2022/7/8 | 2034 | 1600.7 | 1587.9 | 1613.9 |
| 2022/6/1 | 136 | 91.7 | 91.0529 | 92.2739 | 2022/7/9 | 2034 | 1695.5 | 1682.9 | 1708.9 |
| 2022/6/2 | 162 | 101.9 | 101.2 | 102.7 | 2022/7/10 | 2034 | 1795.2 | 1782.7 | 1808.7 |
| 2022/6/3 | 181 | 113.0 | 112.2 | 113.9 | 2022/7/11 | 2034 | 1900.0 | 1886.9 | 1912.6 |
| 2022/6/4 | 181 | 125.0 | 124 | 126 | 2022/7/12 | 2447 | 2010.2 | 1998.2 | 2023.9 |
| 2022/6/5 | 181 | 138.0 | 136.8 | 139.1 | 2022/7/13 | 2447 | 2126.0 | 2113.4 | 2139 |
| 2022/6/6 | 198 | 151.9 | 150.6 | 153.3 | 2022/7/14 | 2447 | 2247.8 | 2234.6 | 2260.7 |
| 2022/6/7 | 225 | 167.0 | 165.5 | 168.6 | 2022/7/15 | 2835 | 2375.8 | 2362.7 | 2389.5 |
| 2022/6/8 | 259 | 183.3 | 181.6 | 185.1 | 2022/7/16 | 2835 | 2510.3 | 2496.8 | 2524.5 |
| 2022/6/9 | 259 | 200.9 | 198.9 | 203 | 2022/7/17 | 2835 | 2651.7 | 2637 | 2666.4 |
| 2022/6/10 | 275 | 219.9 | 217.6 | 222.3 | 2022/7/18 | 2835 | 2774.6 | 2760.3 | 2790.9 |
| 2022/6/11 | 275 | 240.5 | 237.8 | 243.1 | 2022/7/19 | 3125 | 2895.7 | 2880.2 | 2912 |
| 2022/6/12 | 275 | 262.6 | 259.6 | 265.7 | 2022/7/20 | 3125 | 3014.8 | 2998.1 | 3030.9 |
| 2022/6/13 | 275 | 286.6 | 283.1 | 290.1 | 2022/7/21 | 3125 | 3132.0 | 3115 | 3149 |
| 2022/6/14 | 313 | 312.4 | 308.5 | 316.4 | 2022/7/22 | 3126 | 3247.3 | 3230 | 3265.3 |
| 2022/6/15 | 313 | 340.3 | 335.8 | 344.8 | 2022/7/23 | 3126 | 3360.8 | 3342.7 | 3379 |
| 2022/6/16 | 497 | 370.5 | 365.3 | 375.4 | 2022/7/24 | 3126 | 3472.5 | 3453.6 | 3490.7 |
| 2022/6/17 | 498 | 403.0 | 397.1 | 408.6 | 2022/7/25 | 3126 | 3582.4 | 3562.8 | 3600.8 |
| 2022/6/18 | 498 | 435.9 | 429.4 | 442 | 2022/7/26 | 3738 | 3690.5 | 3670.7 | 3709.1 |
| 2022/6/19 | 498 | 470.4 | 463.3 | 477 | 2022/7/27 | 3738 | 3796.9 | 3777.1 | 3816 |
| 2022/6/20 | 521 | 506.7 | 499.1 | 513.8 | 2022/7/28 | 3738 | 3901.5 | 3881.8 | 3920.9 |
| 2022/6/21 | 521 | 544.9 | 536.7 | 552.5 | 2022/7/29 | 4299 | 4004.5 | 3984.5 | 4023.9 |
| 2022/6/22 | 521 | 585.1 | 576.4 | 593.1 | 2022/7/30 | 4300 | 4105.7 | 4085.4 | 4124.9 |
| 2022/6/23 | 736 | 627.4 | 618.1 | 635.9 | 2022/7/31 | 4300 | 4205.4 | 4185.4 | 4224.8 |
| 2022/6/24 | 736 | 671.9 | 662 | 680.8 | 2022/8/1 | 4300 | 4303.4 | 4283.1 | 4322.4 |

CI: credible interval

| Date | Observed cumulated cases | Predicted cumulated cases | 95% CI | | Date | Observed cumulated cases | Predicted cumulated cases | 95% CI | |
|---|---|---|---|---|---|---|---|---|---|
| 2022/8/2 | 4577 | 4399.7 | 4379.8 | 4418.8 | 2022/9/9 | 6884 | 6887.5 | 6865.8 | 6909.5 |
| 2022/8/3 | 4577 | 4494.6 | 4475 | 4513.8 | 2022/9/10 | 6884 | 6905.3 | 6883.4 | 6927.3 |
| 2022/8/4 | 4577 | 4587.8 | 4568.6 | 4607.1 | 2022/9/11 | 6884 | 6921.2 | 6899.1 | 6944.4 |
| 2022/8/5 | 4942 | 4679.5 | 4660.6 | 4698.6 | 2022/9/12 | 6884 | 6935.6 | 6912.9 | 6960.4 |
| 2022/8/6 | 4942 | 4769.7 | 4751.1 | 4788.8 | 2022/9/13 | 6947 | 6948.5 | 6924.7 | 6975 |
| 2022/8/7 | 4942 | 4858.4 | 4840.5 | 4877.8 | 2022/9/14 | 6947 | 6960.1 | 6933.9 | 6987.3 |
| 2022/8/8 | 4942 | 4945.7 | 4927.7 | 4964.5 | 2022/9/15 | 6947 | 6970.5 | 6942.4 | 6999.2 |
| 2022/8/9 | 5162 | 5031.4 | 5014.3 | 5050.3 | 2022/9/16 | 7037 | 6979.9 | 6950.7 | 7010.9 |
| 2022/8/10 | 5162 | 5115.8 | 5098.7 | 5134.4 | 2022/9/17 | 7037 | 6988.3 | 6957.3 | 7021.3 |
| 2022/8/11 | 5162 | 5198.8 | 5181.5 | 5216.7 | 2022/9/18 | 7037 | 6995.9 | 6962.1 | 7029.8 |
| 2022/8/12 | 5719 | 5280.3 | 5262.7 | 5297.7 | 2022/9/19 | 7037 | 7002.8 | 6967.9 | 7039.4 |
| 2022/8/13 | 5719 | 5360.5 | 5343.6 | 5378.3 | 2022/9/20 | 7083 | 7008.9 | 6971.7 | 7046.6 |
| 2022/8/14 | 5719 | 5439.4 | 5422.2 | 5457.1 | 2022/9/21 | 7083 | 7014.4 | 6975.8 | 7054.1 |
| 2022/8/15 | 5719 | 5516.9 | 5499.8 | 5534.8 | 2022/9/22 | 7083 | 7019.4 | 6978.4 | 7060.1 |
| 2022/8/16 | 5792 | 5593.2 | 5575.4 | 5611 | 2022/9/23 | 7083 | 7023.9 | 6981.6 | 7066.4 |
| 2022/8/17 | 5792 | 5668.1 | 5650.1 | 5686.3 | 2022/9/9 | 6884 | 7027.9 | 6984.7 | 7072.5 |
| 2022/8/18 | 5792 | 5741.8 | 5723.8 | 5761 | 2022/9/10 | 6884 | 7031.6 | 6986.8 | 7077.7 |
| 2022/8/19 | 6119 | 5814.2 | 5795.7 | 5834.1 | 2022/9/11 | 6884 | 7034.8 | 6988.6 | 7082.3 |
| 2022/8/20 | 6119 | 5885.4 | 5866.1 | 5905.5 | 2022/9/12 | 6884 | 7037.8 | 6990.5 | 7086.9 |
| 2022/8/21 | 6119 | 5955.4 | 5935.3 | 5976.1 | 2022/9/13 | 6947 | 7040.4 | 6992 | 7091 |
| 2022/8/22 | 6119 | 6024.3 | 6002.3 | 6045 | 2022/9/14 | 6947 | 7042.8 | 6990.2 | 7091.4 |
| 2022/8/23 | 6284 | 6091.9 | 6071.1 | 6115.6 | 2022/9/15 | 6947 | 7045.0 | 6994.9 | 7098.4 |
| 2022/8/24 | 6284 | 6158.4 | 6134.9 | 6181.4 | 2022/9/16 | 7037 | 6887.5 | 6865.8 | 6909.5 |
| 2022/8/25 | 6284 | 6223.8 | 6199.2 | 6247.9 | 2022/9/17 | 7037 | 6905.3 | 6883.4 | 6927.3 |
| 2022/8/26 | 6459 | 6288.0 | 6262.9 | 6314 | 2022/9/18 | 7037 | 6921.2 | 6899.1 | 6944.4 |
| 2022/8/27 | 6459 | 6351.2 | 6324.7 | 6378.3 | 2022/9/19 | 7037 | 6935.6 | 6912.9 | 6960.4 |
| 2022/8/28 | 6459 | 6413.2 | 6386 | 6442.2 | 2022/9/20 | 7083 | 6948.5 | 6924.7 | 6975 |
| 2022/8/29 | 6459 | 6474.2 | 6446.3 | 6505.2 | 2022/9/21 | 7083 | 6960.1 | 6933.9 | 6987.3 |
| 2022/8/30 | 6543 | 6534.2 | 6505.3 | 6567.2 | 2022/9/22 | 7083 | 6970.5 | 6942.4 | 6999.2 |
| 2022/8/31 | 6543 | 6593.1 | 6561.9 | 6626.7 | 2022/9/23 | 7083 | 6979.9 | 6950.7 | 7010.9 |
| 2022/9/1 | 6543 | 6651.0 | 6618.8 | 6686.3 | 2022/9/24 | | 6988.3 | 6957.3 | 7021.3 |
| 2022/9/2 | 6645 | 6692.7 | 6662.5 | 6727.2 | 2022/9/25 | | 6995.9 | 6962.1 | 7029.8 |
| 2022/9/3 | 6645 | 6730.2 | 6699 | 6760.5 | 2022/9/26 | | 7002.8 | 6967.9 | 7039.4 |
| 2022/9/4 | 6645 | 6763.9 | 6735.4 | 6792.6 | 2022/9/27 | | 7008.9 | 6971.7 | 7046.6 |
| 2022/9/5 | 6645 | 6794.2 | 6765.8 | 6819 | 2022/9/28 | | 7014.4 | 6975.8 | 7054.1 |
| 2022/9/6 | 6749 | 6821.4 | 6796.7 | 6846.1 | 2022/9/29 | | 7019.4 | 6978.4 | 7060.1 |
| 2022/9/7 | 6749 | 6845.8 | 6822.4 | 6869.2 | 2022/9/30 | | 7023.9 | 6981.6 | 7066.4 |
| 2022/9/8 | 6749 | 6867.8 | 6846.1 | 6890.9 | | | | | |

CI: credible interval

### (d) Brazil

| Date | Observed cumulated cases | Predicted cumulated cases | 95% CI | | Date | Observed cumulated cases | Predicted cumulated cases | 95% CI | |
|---|---|---|---|---|---|---|---|---|---|
| 2022/6/8 | 1 | 1.0 | | | 2022/7/16 | 347 | 355.8 | 349.0 | 362.6 |
| 2022/6/9 | 1 | 1.5 | 1.5 | 1.5 | 2022/7/17 | 347 | 388.8 | 381.6 | 396.0 |
| 2022/6/10 | 1 | 2.1 | 2.1 | 2.1 | 2022/7/18 | 347 | 424.2 | 416.5 | 431.8 |
| 2022/6/11 | 2 | 2.8 | 2.8 | 2.8 | 2022/7/19 | 448 | 462.1 | 454.1 | 470.2 |
| 2022/6/12 | 3 | 3.5 | 3.5 | 3.6 | 2022/7/20 | 591 | 502.8 | 494.4 | 511.4 |
| 2022/6/13 | 3 | 4.4 | 4.4 | 4.4 | 2022/7/21 | 604 | 546.5 | 537.7 | 555.5 |
| 2022/6/14 | 5 | 5.4 | 5.3 | 5.4 | 2022/7/22 | 694 | 593.4 | 584.2 | 602.8 |
| 2022/6/15 | 5 | 6.5 | 6.4 | 6.5 | 2022/7/23 | 694 | 643.6 | 633.7 | 653.0 |
| 2022/6/16 | 6 | 7.7 | 7.7 | 7.8 | 2022/7/24 | 694 | 697.6 | 687.5 | 707.6 |
| 2022/6/17 | 7 | 9.1 | 9.0 | 9.2 | 2022/7/25 | 809 | 755.4 | 744.5 | 765.2 |
| 2022/6/18 | 7 | 10.7 | 10.6 | 10.8 | 2022/7/26 | 865 | 817.4 | 806.3 | 827.4 |
| 2022/6/19 | 8 | 12.5 | 12.4 | 12.6 | 2022/7/27 | 977 | 884.0 | 873.5 | 895.1 |
| 2022/6/20 | 8 | 14.6 | 14.4 | 14.7 | 2022/7/28 | 1066 | 955.3 | 945.1 | 967.0 |
| 2022/6/21 | 9 | 16.9 | 16.7 | 17.0 | 2022/7/29 | 1259 | 1031.9 | 1021.5 | 1043.7 |
| 2022/6/22 | 11 | 19.5 | 19.3 | 19.7 | 2022/7/30 | 1343 | 1113.9 | 1103.6 | 1125.9 |
| 2022/6/23 | 16 | 22.5 | 22.2 | 22.7 | 2022/7/31 | 1370 | 1202.0 | 1190.4 | 1212.7 |
| 2022/6/24 | 17 | 25.8 | 25.6 | 26.1 | 2022/8/1 | 1475 | 1296.4 | 1285.5 | 1307.6 |
| 2022/6/25 | 19 | 29.6 | 29.3 | 30.0 | 2022/8/2 | 1603 | 1397.6 | 1386.0 | 1408.4 |
| 2022/6/26 | 20 | 34.0 | 33.6 | 34.4 | 2022/8/3 | 1721 | 1506.1 | 1495.0 | 1517.7 |
| 2022/6/27 | 20 | 38.8 | 38.4 | 39.4 | 2022/8/4 | 1860 | 1622.5 | 1611.4 | 1634.2 |
| 2022/6/28 | 21 | 44.4 | 43.8 | 45.0 | 2022/8/5 | 2004 | 1747.2 | 1735.5 | 1759.0 |
| 2022/6/29 | 37 | 50.6 | 50.0 | 51.4 | 2022/8/6 | 2108 | 1881.0 | 1868.3 | 1893.1 |
| 2022/6/30 | 49 | 57.7 | 56.9 | 58.6 | 2022/8/7 | 2131 | 2024.4 | 2011.8 | 2038.2 |
| 2022/7/1 | 64 | 65.7 | 64.8 | 66.8 | 2022/8/8 | 2293 | 2147.6 | 2134.5 | 2162.1 |
| 2022/7/2 | 76 | 74.8 | 73.7 | 76.1 | 2022/8/9 | 2415 | 2270.4 | 2255.7 | 2284.6 |
| 2022/7/3 | 78 | 85.1 | 83.7 | 86.6 | 2022/8/10 | 2458 | 2392.7 | 2377.2 | 2407.5 |
| 2022/7/4 | 80 | 96.8 | 95.1 | 98.5 | 2022/8/11 | 2458 | 2514.5 | 2498.8 | 2530.5 |
| 2022/7/5 | 106 | 110.0 | 108.1 | 112.1 | 2022/8/12 | 2746 | 2635.9 | 2619.3 | 2652.2 |
| 2022/7/6 | 142 | 124.9 | 122.6 | 127.4 | 2022/8/13 | 2848 | 2756.8 | 2739.5 | 2773.6 |
| 2022/7/7 | 172 | 141.9 | 139.1 | 144.8 | 2022/8/14 | 2893 | 2877.2 | 2859.0 | 2893.9 |
| 2022/7/8 | 204 | 161.0 | 157.8 | 164.5 | 2022/8/15 | 2985 | 2997.2 | 2979.0 | 3014.9 |
| 2022/7/9 | 218 | 179.8 | 176.2 | 183.7 | 2022/8/16 | 3183 | 3116.7 | 3097.7 | 3134.5 |
| 2022/7/10 | 218 | 200.0 | 195.9 | 204.3 | 2022/8/17 | 3359 | 3235.7 | 3216.5 | 3254.0 |
| 2022/7/11 | 228 | 221.6 | 217.1 | 226.3 | 2022/8/18 | 3450 | 3354.2 | 3335.6 | 3373.6 |
| 2022/7/12 | 266 | 244.8 | 239.8 | 249.9 | 2022/8/19 | 3655 | 3472.2 | 3453.4 | 3491.8 |
| 2022/7/13 | 308 | 269.7 | 264.2 | 275.2 | 2022/8/20 | 3755 | 3589.8 | 3570.6 | 3609.3 |
| 2022/7/14 | 347 | 296.4 | 290.6 | 302.5 | 2022/8/21 | 3787 | 3706.8 | 3687.4 | 3726.4 |
| 2022/7/15 | 347 | 325.1 | 318.7 | 331.4 | 2022/8/22 | 3896 | 3823.4 | 3804.3 | 3843.3 |

| Date | Observed cumulated cases | Predicted cumulated cases | 95% CI | | Date | Observed cumulated cases | Predicted cumulated cases | 95% CI | |
|---|---|---|---|---|---|---|---|---|---|
| 2022/8/23 | 3984 | 3939.4 | 3919.5 | 3958.7 | 2022/9/30 | | 7959.5 | 7899.1 | 8017.8 |
| 2022/8/24 | 4144 | 4054.9 | 4035.1 | 4074.2 | | | | | |
| 2022/8/25 | 4216 | 4170.0 | 4151.2 | 4190.1 | | | | | |
| 2022/8/26 | 4472 | 4284.5 | 4264.7 | 4303.4 | | | | | |
| 2022/8/27 | 4472 | 4398.5 | 4379.5 | 4417.9 | | | | | |
| 2022/8/28 | 4472 | 4512.0 | 4493.2 | 4531.1 | | | | | |
| 2022/8/29 | 4692 | 4625.0 | 4606.8 | 4644.4 | | | | | |
| 2022/8/30 | 4876 | 4737.5 | 4719.1 | 4756.7 | | | | | |
| 2022/8/31 | 5037 | 4849.5 | 4830.9 | 4868.2 | | | | | |
| 2022/9/1 | 5197 | 4960.9 | 4942.2 | 4979.3 | | | | | |
| 2022/9/2 | 5197 | 5071.8 | 5053.3 | 5090.4 | | | | | |
| 2022/9/3 | 5197 | 5182.2 | 5162.9 | 5200.0 | | | | | |
| 2022/9/4 | 5409 | 5292.1 | 5273.1 | 5310.0 | | | | | |
| 2022/9/5 | 5525 | 5401.4 | 5382.6 | 5419.3 | | | | | |
| 2022/9/6 | 5692 | 5510.2 | 5491.3 | 5528.6 | | | | | |
| 2022/9/7 | 5726 | 5618.5 | 5599.2 | 5637.1 | | | | | |
| 2022/9/8 | 5852 | 5726.3 | 5705.6 | 5744.6 | | | | | |
| 2022/9/9 | 5971 | 5833.5 | 5812.3 | 5852.5 | | | | | |
| 2022/9/10 | 6014 | 5940.1 | 5919.1 | 5960.9 | | | | | |
| 2022/9/11 | 6032 | 6046.3 | 6025.0 | 6068.4 | | | | | |
| 2022/9/12 | 6129 | 6151.9 | 6129.8 | 6175.0 | | | | | |
| 2022/9/13 | 6246 | 6256.9 | 6232.1 | 6279.1 | | | | | |
| 2022/9/14 | 6448 | 6361.5 | 6336.1 | 6385.4 | | | | | |
| 2022/9/15 | 6649 | 6465.4 | 6439.5 | 6491.8 | | | | | |
| 2022/9/16 | 6806 | 6568.9 | 6540.8 | 6596.2 | | | | | |
| 2022/9/17 | 6867 | 6671.7 | 6642.9 | 6701.6 | | | | | |
| 2022/9/18 | 6867 | 6774.1 | 6742.5 | 6804.9 | | | | | |
| 2022/9/19 | 7018 | 6875.9 | 6843.0 | 6908.8 | | | | | |
| 2022/9/20 | 7115 | 6977.1 | 6942.5 | 7012.5 | | | | | |
| 2022/9/21 | 7205 | 7077.8 | 7040.5 | 7114.6 | | | | | |
| 2022/9/22 | 7300 | 7178.0 | 7138.7 | 7217.2 | | | | | |
| 2022/9/23 | 7300 | 7277.6 | 7236.2 | 7318.8 | | | | | |
| 2022/9/24 | | 7376.7 | 7332.5 | 7419.5 | | | | | |
| 2022/9/25 | | 7475.2 | 7428.2 | 7520.2 | | | | | |
| 2022/9/26 | | 7573.1 | 7523.4 | 7620.6 | | | | | |
| 2022/9/27 | | 7670.5 | 7617.7 | 7719.9 | | | | | |
| 2022/9/28 | | 7767.4 | 7712.1 | 7820.1 | | | | | |
| 2022/9/29 | | 7863.7 | 7806.0 | 7919.2 | | | | | |

### (e) United Kingdom

| Date | Observed cumulated cases | Predicted cumulated cases | 95% CI | | Date | Observed cumulated cases | Predicted cumulated cases | 95% CI | |
|---|---|---|---|---|---|---|---|---|---|
| 2022/5/6 | 1 | 1 | | | 2022/6/13 | 470 | 446.7 | 439.0 | 454.2 |
| 2022/5/7 | 1 | 1.6 | 1.6 | 1.6 | 2022/6/14 | 524 | 478.2 | 470.0 | 485.9 |
| 2022/5/8 | 1 | 2.2 | 2.2 | 2.3 | 2022/6/15 | 524 | 510.5 | 502.1 | 518.5 |
| 2022/5/9 | 1 | 3.0 | 3.0 | 3.0 | 2022/6/16 | 574 | 543.8 | 535.5 | 552.5 |
| 2022/5/10 | 1 | 3.9 | 3.9 | 3.9 | 2022/6/17 | 574 | 578.1 | 569.5 | 586.9 |
| 2022/5/11 | 1 | 4.9 | 4.8 | 4.9 | 2022/6/18 | 574 | 613.4 | 604.5 | 622.3 |
| 2022/5/12 | 2 | 6.0 | 6.0 | 6.0 | 2022/6/19 | 574 | 649.6 | 640.5 | 658.6 |
| 2022/5/13 | 3 | 7.3 | 7.3 | 7.3 | 2022/6/20 | 793 | 687.0 | 677.9 | 696.1 |
| 2022/5/14 | 3 | 8.8 | 8.7 | 8.8 | 2022/6/21 | 793 | 725.4 | 716.1 | 734.5 |
| 2022/5/15 | 7 | 10.5 | 10.4 | 10.6 | 2022/6/22 | 793 | 764.9 | 755.6 | 774.0 |
| 2022/5/16 | 7 | 12.4 | 12.4 | 12.5 | 2022/6/23 | 910 | 805.5 | 796.1 | 814.7 |
| 2022/5/17 | 7 | 14.7 | 14.6 | 14.8 | 2022/6/24 | 910 | 847.3 | 837.9 | 856.5 |
| 2022/5/18 | 9 | 17.2 | 17.1 | 17.3 | 2022/6/25 | 910 | 890.3 | 881.6 | 900.2 |
| 2022/5/19 | 9 | 20.1 | 20.0 | 20.3 | 2022/6/26 | 1076 | 934.6 | 925.9 | 944.4 |
| 2022/5/20 | 20 | 23.5 | 23.3 | 23.7 | 2022/6/27 | 1076 | 980.1 | 971.5 | 989.9 |
| 2022/5/21 | 20 | 27.3 | 27.0 | 27.5 | 2022/6/28 | 1076 | 1027.0 | 1018.2 | 1036.5 |
| 2022/5/22 | 20 | 31.7 | 31.4 | 32.0 | 2022/6/29 | 1076 | 1075.1 | 1066.2 | 1084.4 |
| 2022/5/23 | 57 | 36.6 | 36.3 | 37.0 | 2022/6/30 | 1235 | 1124.7 | 1116.0 | 1134.2 |
| 2022/5/24 | 71 | 42.4 | 41.9 | 42.8 | 2022/7/1 | 1235 | 1175.7 | 1166.4 | 1184.8 |
| 2022/5/25 | 78 | 48.9 | 48.3 | 49.5 | 2022/7/2 | 1235 | 1228.1 | 1218.1 | 1237.0 |
| 2022/5/26 | 106 | 56.4 | 55.7 | 57.1 | 2022/7/3 | 1235 | 1282.0 | 1272.0 | 1291.3 |
| 2022/5/27 | 106 | 64.9 | 64.1 | 65.7 | 2022/7/4 | 1351 | 1337.5 | 1327.4 | 1347.6 |
| 2022/5/28 | 106 | 74.7 | 73.7 | 75.7 | 2022/7/5 | 1351 | 1394.5 | 1384.2 | 1405.4 |
| 2022/5/29 | 106 | 85.9 | 84.6 | 87.1 | 2022/7/6 | 1351 | 1446.6 | 1435.9 | 1457.9 |
| 2022/5/30 | 179 | 98.7 | 97.2 | 100.1 | 2022/7/7 | 1552 | 1497.9 | 1486.8 | 1509.7 |
| 2022/5/31 | 190 | 113.3 | 111.5 | 115.0 | 2022/7/8 | 1552 | 1548.5 | 1537.0 | 1560.7 |
| 2022/6/1 | 196 | 130.1 | 127.8 | 132.1 | 2022/7/9 | 1552 | 1598.5 | 1586.3 | 1610.8 |
| 2022/6/2 | 207 | 149.3 | 146.7 | 151.7 | 2022/7/10 | 1552 | 1647.8 | 1635.8 | 1661.0 |
| 2022/6/3 | 226 | 171.2 | 168.0 | 174.1 | 2022/7/11 | 1735 | 1696.4 | 1683.8 | 1709.8 |
| 2022/6/4 | 226 | 196.3 | 192.5 | 199.8 | 2022/7/12 | 1735 | 1744.4 | 1731.0 | 1757.6 |
| 2022/6/5 | 226 | 224.9 | 220.4 | 229.1 | 2022/7/13 | 1735 | 1791.8 | 1778.3 | 1805.3 |
| 2022/6/6 | 302 | 250.0 | 245.1 | 254.7 | 2022/7/14 | 1856 | 1838.5 | 1824.9 | 1852.4 |
| 2022/6/7 | 321 | 275.7 | 270.3 | 280.8 | 2022/7/15 | 1856 | 1884.6 | 1870.7 | 1898.6 |
| 2022/6/8 | 321 | 302.2 | 296.4 | 307.8 | 2022/7/16 | 1856 | 1930.1 | 1916.0 | 1944.2 |
| 2022/6/9 | 366 | 329.5 | 323.2 | 335.5 | 2022/7/17 | 1856 | 1975.0 | 1960.8 | 1989.4 |
| 2022/6/10 | 366 | 357.5 | 350.9 | 364.0 | 2022/7/18 | 2137 | 2019.2 | 2005.0 | 2033.7 |
| 2022/6/11 | 366 | 386.4 | 379.4 | 393.2 | 2022/7/19 | 2137 | 2062.9 | 2048.5 | 2077.2 |
| 2022/6/12 | 470 | 416.1 | 408.8 | 423.3 | 2022/7/20 | 2137 | 2106.0 | 2091.4 | 2120.2 |

| Date | Observed cumulated cases | Predicted cumulated cases | 95% CI | | Date | Observed cumulated cases | Predicted cumulated cases | 95% CI | |
|---|---|---|---|---|---|---|---|---|---|
| 2022/7/21 | 2208 | 2148.5 | 2134.3 | 2163.2 | 2022/8/28 | 3340 | 3397.5 | 3366.5 | 3419.0 |
| 2022/7/22 | 2208 | 2190.4 | 2176.5 | 2205.4 | 2022/8/29 | 3413 | 3422.2 | 3392.1 | 3446.6 |
| 2022/7/23 | 2208 | 2231.8 | 2218.1 | 2246.8 | 2022/8/30 | 3413 | 3446.7 | 3414.2 | 3470.8 |
| 2022/7/24 | 2208 | 2272.6 | 2258.8 | 2287.3 | 2022/8/31 | 3413 | 3470.7 | 3437.9 | 3496.5 |
| 2022/7/25 | 2497 | 2312.8 | 2299.2 | 2327.7 | 2022/9/1 | 3413 | 3494.5 | 3460.5 | 3521.5 |
| 2022/7/26 | 2497 | 2352.5 | 2338.9 | 2367.2 | 2022/9/2 | 3413 | 3504.1 | 3472.8 | 3526.6 |
| 2022/7/27 | 2497 | 2391.7 | 2378.0 | 2405.9 | 2022/9/3 | 3413 | 3511.9 | 3485.6 | 3532.2 |
| 2022/7/28 | 2546 | 2430.3 | 2417.0 | 2444.7 | 2022/9/4 | 3413 | 3518.3 | 3496.6 | 3537.1 |
| 2022/7/29 | 2546 | 2468.4 | 2455.2 | 2482.6 | 2022/9/5 | 3484 | 3523.4 | 3504.0 | 3540.3 |
| 2022/7/30 | 2546 | 2506.0 | 2492.5 | 2519.5 | 2022/9/6 | 3484 | 3527.6 | 3509.7 | 3543.8 |
| 2022/7/31 | 2546 | 2543.1 | 2530.3 | 2557.1 | 2022/9/7 | 3484 | 3531.0 | 3514.8 | 3548.8 |
| 2022/8/1 | 2759 | 2579.6 | 2566.0 | 2592.5 | 2022/9/8 | 3484 | 3533.8 | 3516.9 | 3552.4 |
| 2022/8/2 | 2759 | 2615.7 | 2602.4 | 2628.6 | 2022/9/9 | 3484 | 3536.1 | 3518.3 | 3556.0 |
| 2022/8/3 | 2759 | 2651.3 | 2638.8 | 2664.9 | 2022/9/10 | 3484 | 3538.0 | 3518.0 | 3558.5 |
| 2022/8/4 | 2859 | 2686.4 | 2673.2 | 2699.2 | 2022/9/11 | 3484 | 3539.6 | 3519.9 | 3563.8 |
| 2022/8/5 | 2859 | 2721.0 | 2707.9 | 2733.6 | 2022/9/12 | 3552 | 3540.9 | 3516.2 | 3563.0 |
| 2022/8/6 | 2859 | 2755.1 | 2741.9 | 2767.6 | 2022/9/13 | 3552 | 3542.0 | 3517.8 | 3567.6 |
| 2022/8/7 | 2859 | 2788.8 | 2775.6 | 2801.5 | 2022/9/14 | 3552 | 3542.9 | 3517.0 | 3569.4 |
| 2022/8/8 | 3017 | 2822.0 | 2808.7 | 2834.7 | 2022/9/15 | 3552 | 3543.7 | 3517.2 | 3572.1 |
| 2022/8/9 | 3017 | 2854.7 | 2841.1 | 2867.3 | 2022/9/16 | 3552 | 3544.3 | 3516.9 | 3574.1 |
| 2022/8/10 | 3017 | 2887.0 | 2873.0 | 2900.0 | 2022/9/17 | 3552 | 3544.9 | 3517.7 | 3576.9 |
| 2022/8/11 | 3017 | 2918.8 | 2904.3 | 2932.1 | 2022/9/18 | 3552 | 3545.3 | 3517.9 | 3578.8 |
| 2022/8/12 | 3017 | 2950.2 | 2934.9 | 2963.6 | 2022/9/19 | 3552 | 3545.7 | 3517.3 | 3579.8 |
| 2022/8/13 | 3017 | 2981.2 | 2965.7 | 2995.2 | 2022/9/20 | 3585 | 3546.0 | 3518.0 | 3581.9 |
| 2022/8/14 | 3017 | 3011.8 | 2996.1 | 3026.4 | 2022/9/21 | 3585 | 3546.3 | 3518.1 | 3583.1 |
| 2022/8/15 | 3195 | 3041.9 | 3025.5 | 3056.9 | 2022/9/22 | 3585 | 3546.5 | 3518.5 | 3584.5 |
| 2022/8/16 | 3195 | 3071.6 | 3054.5 | 3087.1 | 2022/9/23 | 3585 | 3546.7 | 3518.5 | 3585.5 |
| 2022/8/17 | 3195 | 3100.9 | 3082.3 | 3115.9 | 2022/9/24 | | 3546.9 | 3518.2 | 3586.1 |
| 2022/8/18 | 3195 | 3129.8 | 3111.7 | 3146.8 | 2022/9/25 | | 3547.1 | 3518.2 | 3586.9 |
| 2022/8/19 | 3195 | 3158.3 | 3138.3 | 3174.7 | 2022/9/26 | | 3547.2 | 3517.4 | 3586.7 |
| 2022/8/20 | 3195 | 3186.4 | 3165.9 | 3203.6 | 2022/9/27 | | 3547.3 | 3517.4 | 3587.3 |
| 2022/8/21 | 3195 | 3214.1 | 3192.3 | 3231.8 | 2022/9/28 | | 3547.4 | 3517.4 | 3587.7 |
| 2022/8/22 | 3340 | 3241.4 | 3217.8 | 3258.7 | 2022/9/29 | | 3547.5 | 3517.4 | 3588.2 |
| 2022/8/23 | 3340 | 3268.3 | 3244.2 | 3286.9 | 2022/9/30 | | 3547.5 | 3517.1 | 3588.2 |
| 2022/8/24 | 3340 | 3294.9 | 3268.7 | 3313.4 | | | | | |
| 2022/8/25 | 3340 | 3321.1 | 3294.3 | 3340.8 | | | | | |
| 2022/8/26 | 3340 | 3346.9 | 3318.5 | 3367.0 | | | | | |
| 2022/8/27 | 3340 | 3372.4 | 3342.6 | 3393.1 | | | | | |

**Table A2. Estimated results on parameters for Monkeypox by using the deterministic SIR model.**

| Country/Areas | The periods | The deterministic SIR model | |
|---|---|---|---|
| | | **β** | **$R_0/R_e$** |
| **The Globe** | January 31 ~ April 30 | 0.132 | 1.119 |
| | May 1 ~ May 31 | 0.265 | 2.251 |
| | June 1 ~ June 30 | 0.180 | 1.532 |
| | July 1 ~ July 31 | 0.149 | 1.271 |
| | August 1 ~ August 31 | 0.136 | 1.153 |
| | September 1 ~September 23 | 0.096 | 0.817 |
| **United States** | May 18 ~ June 30 | 0.245 | 2.086 |
| | July 1 ~ July 30 | 0.184 | 1.568 |
| | August 1 ~ August 31 | 0.145 | 1.229 |
| | September 1 ~September 23 | 0.080 | 0.677 |
| **Spain** | May 18 ~ June 16 | 0.242 | 2.057 |
| | June 17 ~ July 16 | 0.15318 | 1.302 |
| | July 17 ~ August 31 | 0.11767 | 1.000 |
| | September 1 ~September 23 | 0.0523 | 0.445 |
| **Brazil** | June 8 ~ July 7 | 0.276 | 2.346 |
| | July 8 ~ August 6 | 0.1882 | 1.600 |
| | August 7 ~ August 31 | 0.1271 | 1.080 |
| | September 1 ~September 23 | 0.1077 | 0.915 |
| **United Kingdom** | June 8 ~ July 7 | 0.286 | 2.432 |
| | July 8 ~ August 6 | 0.154 | 1.312 |
| | August 7 ~ August 31 | 0.113 | 0.964 |
| | September 1 ~September 23 | 0.055 | 0.470 |

$R_0$: basic reproductive number; $R_e$: effective reproductive number; SIR model: *Susceptible-Infected-Recovered* model.

**Table A3. The observed cumulated monkeypox cases and simulated cumulated cases by the SIR deterministic model**

**(a) The global monkeypox cases**

| Date | Observed cumulated cases | Simulated cumulated cases | Date | Observed cumulated cases | Simulated cumulated cases |
|---|---|---|---|---|---|
| 2022/1/31 | 3 | 1 | 2022/5/8 | 37 | 42 |
| 2022/2/16 | 3 | 3 | 2022/5/9 | 37 | 46 |
| 2022/2/17 | 6 | 4 | 2022/5/10 | 37 | 50 |
| 2022/2/27 | 6 | 5 | 2022/5/11 | 37 | 55 |
| 2022/2/28 | 7 | 6 | 2022/5/12 | 38 | 61 |
| 2022/3/3 | 7 | 6 | 2022/5/13 | 39 | 67 |
| 2022/3/4 | 9 | 7 | 2022/5/14 | 39 | 75 |
| 2022/3/30 | 9 | 13 | 2022/5/15 | 43 | 84 |
| 2022/3/31 | 15 | 14 | 2022/5/16 | 43 | 94 |
| 2022/4/9 | 15 | 17 | 2022/5/17 | 46 | 106 |
| 2022/4/10 | 19 | 17 | 2022/5/18 | 67 | 120 |
| 2022/4/11 | 19 | 18 | 2022/5/19 | 86 | 136 |
| 2022/4/12 | 21 | 18 | 2022/5/20 | 131 | 155 |
| 2022/4/13 | 21 | 19 | 2022/5/21 | 147 | 176 |
| 2022/4/14 | 21 | 19 | 2022/5/22 | 147 | 201 |
| 2022/4/15 | 21 | 19 | 2022/5/23 | 220 | 230 |
| 2022/4/16 | 21 | 20 | 2022/5/24 | 256 | 263 |
| 2022/4/17 | 21 | 20 | 2022/5/25 | 300 | 302 |
| 2022/4/18 | 21 | 21 | 2022/5/26 | 388 | 347 |
| 2022/4/19 | 21 | 21 | 2022/5/27 | 440 | 399 |
| 2022/4/20 | 21 | 22 | 2022/5/28 | 457 | 460 |
| 2022/4/21 | 21 | 22 | 2022/5/29 | 477 | 529 |
| 2022/4/22 | 21 | 22 | 2022/5/30 | 600 | 611 |
| 2022/4/23 | 21 | 23 | 2022/5/31 | 665 | 671 |
| 2022/4/24 | 21 | 23 | 2022/6/1 | 749 | 736 |
| 2022/4/25 | 21 | 24 | 2022/6/2 | 874 | 805 |
| 2022/4/26 | 21 | 24 | 2022/6/3 | 968 | 879 |
| 2022/4/27 | 21 | 25 | 2022/6/4 | 976 | 957 |
| 2022/4/28 | 21 | 25 | 2022/6/5 | 1083 | 1040 |
| 2022/4/29 | 21 | 26 | 2022/6/6 | 1198 | 1129 |
| 2022/4/30 | 26 | 26 | 2022/6/7 | 1348 | 1224 |
| 2022/5/1 | 26 | 27 | 2022/6/8 | 1445 | 1324 |
| 2022/5/2 | 26 | 29 | 2022/6/9 | 1591 | 1432 |
| 2022/5/3 | 26 | 30 | 2022/6/10 | 1634 | 1546 |
| 2022/5/4 | 26 | 32 | 2022/6/11 | 1643 | 1667 |
| 2022/5/5 | 26 | 34 | 2022/6/12 | 1757 | 1796 |
| 2022/5/6 | 27 | 36 | 2022/6/13 | 1817 | 1934 |
| 2022/5/7 | 27 | 39 | 2022/6/14 | 2061 | 2080 |

| Date | Observed cumulated cases | Simulated cumulated cases | Date | Observed cumulated cases | Simulated cumulated cases |
|---|---|---|---|---|---|
| 2022/6/15 | 2208 | 2235 | 2022/7/23 | 17016 | 17330 |
| 2022/6/16 | 2584 | 2401 | 2022/7/24 | 17050 | 18006 |
| 2022/6/17 | 2778 | 2578 | 2022/7/25 | 18336 | 18702 |
| 2022/6/18 | 2783 | 2765 | 2022/7/26 | 19872 | 19416 |
| 2022/6/19 | 2790 | 2965 | 2022/7/27 | 21025 | 20150 |
| 2022/6/20 | 3084 | 3177 | 2022/7/28 | 21867 | 20905 |
| 2022/6/21 | 3366 | 3403 | 2022/7/29 | 23087 | 21681 |
| 2022/6/22 | 3540 | 3643 | 2022/7/30 | 23172 | 22480 |
| 2022/6/23 | 4098 | 3899 | 2022/7/31 | 23303 | 23306 |
| 2022/6/24 | 4303 | 4172 | 2022/8/1 | 24521 | 24054 |
| 2022/6/25 | 4315 | 4460 | 2022/8/2 | 25963 | 24824 |
| 2022/6/26 | 4511 | 4768 | 2022/8/3 | 26616 | 25605 |
| 2022/6/27 | 4898 | 5096 | 2022/8/4 | 27740 | 26397 |
| 2022/6/28 | 5246 | 5444 | 2022/8/5 | 28907 | 27199 |
| 2022/6/29 | 5399 | 5813 | 2022/8/6 | 29021 | 28013 |
| 2022/6/30 | 6206 | 6208 | 2022/8/7 | 29193 | 28838 |
| 2022/7/1 | 6475 | 6545 | 2022/8/8 | 31178 | 29674 |
| 2022/7/2 | 6519 | 6895 | 2022/8/9 | 32613 | 30521 |
| 2022/7/3 | 6559 | 7258 | 2022/8/10 | 33766 | 31379 |
| 2022/7/4 | 7053 | 7631 | 2022/8/11 | 34451 | 32248 |
| 2022/7/5 | 7500 | 8017 | 2022/8/12 | 35863 | 33128 |
| 2022/7/6 | 7732 | 8413 | 2022/8/13 | 36004 | 34020 |
| 2022/7/7 | 8426 | 8821 | 2022/8/14 | 36172 | 34923 |
| 2022/7/8 | 9486 | 9243 | 2022/8/15 | 37646 | 35838 |
| 2022/7/9 | 9608 | 9678 | 2022/8/16 | 38965 | 36763 |
| 2022/7/10 | 9635 | 10126 | 2022/8/17 | 40147 | 37700 |
| 2022/7/11 | 10226 | 10588 | 2022/8/18 | 41192 | 38647 |
| 2022/7/12 | 11098 | 11064 | 2022/8/19 | 41964 | 39606 |
| 2022/7/13 | 11516 | 11552 | 2022/8/20 | 42130 | 40577 |
| 2022/7/14 | 12127 | 12056 | 2022/8/21 | 42238 | 41558 |
| 2022/7/15 | 13015 | 12576 | 2022/8/22 | 44301 | 42551 |
| 2022/7/16 | 13031 | 13112 | 2022/8/23 | 45752 | 43554 |
| 2022/7/17 | 13069 | 13664 | 2022/8/24 | 46801 | 44569 |
| 2022/7/18 | 13886 | 14232 | 2022/8/25 | 47438 | 45595 |
| 2022/7/19 | 15081 | 14816 | 2022/8/26 | 48504 | 46633 |
| 2022/7/20 | 15655 | 15416 | 2022/8/27 | 48563 | 47681 |
| 2022/7/21 | 16293 | 16035 | 2022/8/28 | 48616 | 48740 |
| 2022/7/22 | 17013 | 16673 | 2022/8/29 | 50343 | 49810 |

| Date | Observed cumulated cases | Simulated cumulated cases | Date | Observed cumulated cases | Simulated cumulated cases |
|---|---|---|---|---|---|
| 2022/8/30 | 51110 | 50891 | | | |
| 2022/8/31 | 51982 | 51988 | | | |
| 2022/9/1 | 52941 | 52744 | | | |
| 2022/9/2 | 53629 | 53490 | | | |
| 2022/9/3 | 53678 | 54216 | | | |
| 2022/9/4 | 53936 | 54920 | | | |
| 2022/9/5 | 54939 | 55606 | | | |
| 2022/9/6 | 56138 | 56274 | | | |
| 2022/9/7 | 56852 | 56925 | | | |
| 2022/9/8 | 57490 | 57560 | | | |
| 2022/9/9 | 58211 | 58177 | | | |
| 2022/9/10 | 58322 | 58778 | | | |
| 2022/9/11 | 58392 | 59362 | | | |
| 2022/9/12 | 59351 | 59928 | | | |
| 2022/9/13 | 60337 | 60477 | | | |
| 2022/9/14 | 60784 | 61011 | | | |
| 2022/9/15 | 61581 | 61531 | | | |
| 2022/9/16 | 62324 | 62038 | | | |
| 2022/9/17 | 62399 | 62531 | | | |
| 2022/9/18 | 62403 | 63012 | | | |
| 2022/9/19 | 63758 | 63479 | | | |
| 2022/9/20 | 64418 | 63933 | | | |
| 2022/9/21 | 64904 | 64373 | | | |
| 2022/9/22 | 65215 | 64800 | | | |
| 2022/9/23 | 65215 | 65214 | | | |

### (b) United States

| Date | Observed cumulated cases | Simulated cumulated cases | Date | Observed cumulated cases | Simulated cumulated cases |
|---|---|---|---|---|---|
| 2022/5/18 | 1 | 1.0 | 2022/6/25 | 243 | 247.7 |
| 2022/5/19 | 2 | 1.3 | 2022/6/26 | 252 | 281.6 |
| 2022/5/20 | 2 | 1.6 | 2022/6/27 | 315 | 320.2 |
| 2022/5/21 | 2 | 1.9 | 2022/6/28 | 359 | 364.0 |
| 2022/5/22 | 2 | 2.3 | 2022/6/29 | 409 | 413.8 |
| 2022/5/23 | 2 | 2.7 | 2022/6/30 | 470 | 470.5 |
| 2022/5/24 | 4 | 3.2 | 2022/7/1 | 543 | 517.0 |
| 2022/5/25 | 6 | 3.8 | 2022/7/2 | 547 | 567.0 |
| 2022/5/26 | 11 | 4.4 | 2022/7/3 | 553 | 620.0 |
| 2022/5/27 | 15 | 5.2 | 2022/7/4 | 563 | 677.0 |
| 2022/5/28 | 16 | 6.0 | 2022/7/5 | 709 | 739.0 |
| 2022/5/29 | 17 | 6.9 | 2022/7/6 | 749 | 804.0 |
| 2022/5/30 | 17 | 8.0 | 2022/7/7 | 883 | 874.0 |
| 2022/5/31 | 21 | 9.2 | 2022/7/8 | 950 | 949.0 |
| 2022/6/1 | 22 | 10.6 | 2022/7/9 | 960 | 1028.0 |
| 2022/6/2 | 26 | 12.2 | 2022/7/10 | 964 | 1114.0 |
| 2022/6/3 | 31 | 14.0 | 2022/7/11 | 1058 | 1205.0 |
| 2022/6/4 | 32 | 16.0 | 2022/7/12 | 1122 | 1303.0 |
| 2022/6/5 | 32 | 18.3 | 2022/7/13 | 1245 | 1407.0 |
| 2022/6/6 | 34 | 20.9 | 2022/7/14 | 1426 | 1519.0 |
| 2022/6/7 | 40 | 23.9 | 2022/7/15 | 1767 | 1638.0 |
| 2022/6/8 | 45 | 27.3 | 2022/7/16 | 1767 | 1766.0 |
| 2022/6/9 | 50 | 31.1 | 2022/7/17 | 1767 | 1903.0 |
| 2022/6/10 | 53 | 35.5 | 2022/7/18 | 1995 | 2049.0 |
| 2022/6/11 | 55 | 40.5 | 2022/7/19 | 2114 | 2204.0 |
| 2022/6/12 | 57 | 46.1 | 2022/7/20 | 2323 | 2371.0 |
| 2022/6/13 | 75 | 52.6 | 2022/7/21 | 2503 | 2550.0 |
| 2022/6/14 | 83 | 59.9 | 2022/7/22 | 2875 | 2740.0 |
| 2022/6/15 | 96 | 68.2 | 2022/7/23 | 2875 | 2944.0 |
| 2022/6/16 | 117 | 77.6 | 2022/7/24 | 2875 | 3163.0 |
| 2022/6/17 | 128 | 88.3 | 2022/7/25 | 3457 | 3396.0 |
| 2022/6/18 | 132 | 100.5 | 2022/7/26 | 3768 | 3645.0 |
| 2022/6/19 | 133 | 114.4 | 2022/7/27 | 4630 | 3912.0 |
| 2022/6/20 | 137 | 130.1 | 2022/7/28 | 4895 | 4197.0 |
| 2022/6/21 | 175 | 148.0 | 2022/7/29 | 5175 | 4501.0 |
| 2022/6/22 | 196 | 168.4 | 2022/7/30 | 5175 | 4826.0 |
| 2022/6/23 | 218 | 191.5 | 2022/7/31 | 5175 | 5174.0 |
| 2022/6/24 | 237 | 217.8 | 2022/8/1 | 5792 | 5466.0 |

| Date | Observed cumulated cases | Simulated cumulated cases | Date | Observed cumulated cases | Simulated cumulated cases |
|---|---|---|---|---|---|
| 2022/8/2 | 6308 | 5760.0 | 2022/9/9 | 21761 | 21626.0 |
| 2022/8/3 | 6599 | 6062.0 | 2022/9/10 | 21761 | 21876.0 |
| 2022/8/4 | 7084 | 6372.0 | 2022/9/11 | 21761 | 22118.0 |
| 2022/8/5 | 7490 | 6691.0 | 2022/9/12 | 21834 | 22351.0 |
| 2022/8/6 | 7491 | 7018.0 | 2022/9/13 | 22480 | 22575.0 |
| 2022/8/7 | 7491 | 7355.0 | 2022/9/14 | 22619 | 22791.0 |
| 2022/8/8 | 8902 | 7702.0 | 2022/9/15 | 22958 | 22999.0 |
| 2022/8/9 | 9461 | 8058.0 | 2022/9/16 | 23339 | 23198.0 |
| 2022/8/10 | 10361 | 8423.0 | 2022/9/17 | 23339 | 23390.0 |
| 2022/8/11 | 10727 | 8798.0 | 2022/9/18 | 23339 | 23575.0 |
| 2022/8/12 | 11131 | 9183.0 | 2022/9/19 | 23730 | 23754.0 |
| 2022/8/13 | 11131 | 9578.0 | 2022/9/20 | 24041 | 23926.0 |
| 2022/8/14 | 11131 | 9985.0 | 2022/9/21 | 24198 | 24091.0 |
| 2022/8/15 | 11844 | 10404.0 | 2022/9/22 | 24403 | 24250.0 |
| 2022/8/16 | 12636 | 10834.0 | 2022/9/23 | 24403 | 24402.0 |
| 2022/8/17 | 13450 | 11276.0 | 2022/9/9 | 21761 | 21626.0 |
| 2022/8/18 | 14049 | 11729.0 | 2022/9/10 | 21761 | 21876.0 |
| 2022/8/19 | 14049 | 12193.0 | 2022/9/11 | 21761 | 22118.0 |
| 2022/8/20 | 14049 | 12670.0 | 2022/9/12 | 21834 | 22351.0 |
| 2022/8/21 | 14049 | 13162.0 | 2022/9/13 | 22480 | 22575.0 |
| 2022/8/22 | 15357 | 13668.0 | 2022/9/14 | 22619 | 22791.0 |
| 2022/8/23 | 15832 | 14187.0 | 2022/9/15 | 22958 | 22999.0 |
| 2022/8/24 | 16514 | 14721.0 | 2022/9/16 | 23339 | 23198.0 |
| 2022/8/25 | 16837 | 15268.0 | 2022/9/17 | 23339 | 23390.0 |
| 2022/8/26 | 17336 | 15830.0 | 2022/9/18 | 23339 | 23575.0 |
| 2022/8/27 | 17336 | 16407.0 | 2022/9/19 | 23730 | 23754.0 |
| 2022/8/28 | 17336 | 16999.0 | 2022/9/20 | 24041 | 23926.0 |
| 2022/8/29 | 17986 | 17608.0 | 2022/9/21 | 24198 | 24091.0 |
| 2022/8/30 | 18298 | 18233.0 | 2022/9/22 | 24403 | 24250.0 |
| 2022/8/31 | 18879 | 18876.0 | 2022/9/23 | 24403 | 24402.0 |
| 2022/9/1 | 19355 | 19236.0 | | | |
| 2022/9/2 | 19852 | 19575.0 | | | |
| 2022/9/3 | 19852 | 19902.0 | | | |
| 2022/9/4 | 19852 | 20216.0 | | | |
| 2022/9/5 | 19852 | 20520.0 | | | |
| 2022/9/6 | 20608 | 20813.0 | | | |
| 2022/9/7 | 21148 | 21095.0 | | | |
| 2022/9/8 | 21374 | 21366.0 | | | |

### (c) Spain

| Date | Observed cumulated cases | Simulated cumulated cases | Date | Observed cumulated cases | Simulated cumulated cases |
|---|---|---|---|---|---|
| 2022/5/18 | 7 | 7.0 | 2022/6/25 | 736 | 919.4 |
| 2022/5/19 | 7 | 8.8 | 2022/6/26 | 736 | 975.2 |
| 2022/5/20 | 30 | 10.8 | 2022/6/27 | 800 | 1032.9 |
| 2022/5/21 | 40 | 13.2 | 2022/6/28 | 800 | 1092.5 |
| 2022/5/22 | 40 | 15.8 | 2022/6/29 | 800 | 1154.4 |
| 2022/5/23 | 41 | 18.8 | 2022/6/30 | 1196 | 1218.6 |
| 2022/5/24 | 45 | 22.1 | 2022/7/1 | 1196 | 1285.1 |
| 2022/5/25 | 53 | 25.9 | 2022/7/2 | 1196 | 1354.0 |
| 2022/5/26 | 78 | 30.2 | 2022/7/3 | 1196 | 1425.2 |
| 2022/5/27 | 99 | 35.1 | 2022/7/4 | 1258 | 1498.9 |
| 2022/5/28 | 99 | 40.7 | 2022/7/5 | 1258 | 1575.5 |
| 2022/5/29 | 108 | 46.9 | 2022/7/6 | 1258 | 1655.0 |
| 2022/5/30 | 116 | 54.1 | 2022/7/7 | 1258 | 1737.2 |
| 2022/5/31 | 136 | 62.1 | 2022/7/8 | 2034 | 1822.3 |
| 2022/6/1 | 136 | 71.2 | 2022/7/9 | 2034 | 1910.1 |
| 2022/6/2 | 162 | 81.6 | 2022/7/10 | 2034 | 2001.4 |
| 2022/6/3 | 181 | 93.2 | 2022/7/11 | 2034 | 2096.0 |
| 2022/6/4 | 181 | 106.5 | 2022/7/12 | 2447 | 2194.2 |
| 2022/6/5 | 181 | 121.5 | 2022/7/13 | 2447 | 2295.8 |
| 2022/6/6 | 198 | 138.5 | 2022/7/14 | 2447 | 2400.8 |
| 2022/6/7 | 225 | 157.8 | 2022/7/15 | 2835 | 2509.5 |
| 2022/6/8 | 259 | 179.6 | 2022/7/16 | 2835 | 2622.0 |
| 2022/6/9 | 259 | 204.3 | 2022/7/17 | 2835 | 2711.7 |
| 2022/6/10 | 275 | 232.3 | 2022/7/18 | 2835 | 2799.8 |
| 2022/6/11 | 275 | 264.0 | 2022/7/19 | 3125 | 2887.9 |
| 2022/6/12 | 275 | 299.8 | 2022/7/20 | 3125 | 2975.9 |
| 2022/6/13 | 275 | 340.5 | 2022/7/21 | 3125 | 3063.9 |
| 2022/6/14 | 313 | 386.5 | 2022/7/22 | 3126 | 3151.9 |
| 2022/6/15 | 313 | 438.7 | 2022/7/23 | 3126 | 3239.8 |
| 2022/6/16 | 497 | 497.8 | 2022/7/24 | 3126 | 3327.7 |
| 2022/6/17 | 498 | 538.5 | 2022/7/25 | 3126 | 3415.6 |
| 2022/6/18 | 498 | 580.4 | 2022/7/26 | 3738 | 3503.4 |
| 2022/6/19 | 498 | 623.8 | 2022/7/27 | 3738 | 3591.1 |
| 2022/6/20 | 521 | 668.8 | 2022/7/28 | 3738 | 3678.8 |
| 2022/6/21 | 521 | 715.5 | 2022/7/29 | 4299 | 3766.5 |
| 2022/6/22 | 521 | 763.8 | 2022/7/30 | 4300 | 3854.1 |
| 2022/6/23 | 736 | 813.8 | 2022/7/31 | 4300 | 3941.7 |
| 2022/6/24 | 736 | 865.6 | 2022/8/1 | 4300 | 4029.2 |

| Date | Observed cumulated cases | Simulated cumulated cases | Date | Observed cumulated cases | Simulated cumulated cases |
|---|---|---|---|---|---|
| 2022/8/2 | 4577 | 4116.7 | 2022/9/9 | 6884 | 6890.4 |
| 2022/8/3 | 4577 | 4204.1 | 2022/9/10 | 6884 | 6910.8 |
| 2022/8/4 | 4577 | 4291.5 | 2022/9/11 | 6884 | 6929.9 |
| 2022/8/5 | 4942 | 4378.9 | 2022/9/12 | 6884 | 6947.9 |
| 2022/8/6 | 4942 | 4466.2 | 2022/9/13 | 6947 | 6964.7 |
| 2022/8/7 | 4942 | 4553.4 | 2022/9/14 | 6947 | 6980.4 |
| 2022/8/8 | 4942 | 4640.6 | 2022/9/15 | 6947 | 6995.1 |
| 2022/8/9 | 5162 | 4727.8 | 2022/9/16 | 7037 | 7009.0 |
| 2022/8/10 | 5162 | 4814.8 | 2022/9/17 | 7037 | 7021.9 |
| 2022/8/11 | 5162 | 4901.8 | 2022/9/18 | 7037 | 7033.9 |
| 2022/8/12 | 5719 | 4988.8 | 2022/9/19 | 7037 | 7045.2 |
| 2022/8/13 | 5719 | 5075.7 | 2022/9/20 | 7083 | 7055.9 |
| 2022/8/14 | 5719 | 5162.6 | 2022/9/21 | 7083 | 7065.8 |
| 2022/8/15 | 5719 | 5249.4 | 2022/9/22 | 7083 | 7075.0 |
| 2022/8/16 | 5792 | 5336.1 | 2022/9/23 | 7083 | 7083.7 |
| 2022/8/17 | 5792 | 5422.8 | 2022/9/9 | 6884 | 6890.4 |
| 2022/8/18 | 5792 | 5509.4 | 2022/9/10 | 6884 | 6910.8 |
| 2022/8/19 | 6119 | 5596.0 | 2022/9/11 | 6884 | 6929.9 |
| 2022/8/20 | 6119 | 5682.5 | 2022/9/12 | 6884 | 6947.9 |
| 2022/8/21 | 6119 | 5768.9 | 2022/9/13 | 6947 | 6964.7 |
| 2022/8/22 | 6119 | 5855.3 | 2022/9/14 | 6947 | 6980.4 |
| 2022/8/23 | 6284 | 5941.6 | 2022/9/15 | 6947 | 6995.1 |
| 2022/8/24 | 6284 | 6027.8 | 2022/9/16 | 7037 | 7009.0 |
| 2022/8/25 | 6284 | 6114.0 | 2022/9/17 | 7037 | 7021.9 |
| 2022/8/26 | 6459 | 6200.0 | 2022/9/18 | 7037 | 7033.9 |
| 2022/8/27 | 6459 | 6286.1 | 2022/9/19 | 7037 | 7045.2 |
| 2022/8/28 | 6459 | 6372.0 | 2022/9/20 | 7083 | 7055.9 |
| 2022/8/29 | 6459 | 6457.9 | 2022/9/21 | 7083 | 7065.8 |
| 2022/8/30 | 6543 | 6543.8 | 2022/9/22 | 7083 | 7075.0 |
| 2022/8/31 | 6543 | 6629.5 | 2022/9/23 | 7083 | 7083.7 |
| 2022/9/1 | 6543 | 6667.7 | | | |
| 2022/9/2 | 6645 | 6702.3 | | | |
| 2022/9/3 | 6645 | 6734.6 | | | |
| 2022/9/4 | 6645 | 6765.0 | | | |
| 2022/9/5 | 6645 | 6793.6 | | | |
| 2022/9/6 | 6749 | 6820.2 | | | |
| 2022/9/7 | 6749 | 6845.1 | | | |
| 2022/9/8 | 6749 | 6868.4 | | | |

**(d) Brazil**

| Date | Observed cumulated cases | Simulated cumulated cases | Date | Observed cumulated cases | Simulated cumulated cases |
|---|---|---|---|---|---|
| 2022/6/8 | 1 | 1 | 2022/7/16 | 347 | 407.9 |
| 2022/6/9 | 1 | 1.3 | 2022/7/17 | 347 | 444.6 |
| 2022/6/10 | 1 | 1.6 | 2022/7/18 | 347 | 483.8 |
| 2022/6/11 | 2 | 2.1 | 2022/7/19 | 448 | 526 |
| 2022/6/12 | 3 | 2.5 | 2022/7/20 | 591 | 571.3 |
| 2022/6/13 | 3 | 3.1 | 2022/7/21 | 604 | 619.7 |
| 2022/6/14 | 5 | 3.8 | 2022/7/22 | 694 | 671.9 |
| 2022/6/15 | 5 | 4.5 | 2022/7/23 | 694 | 727.9 |
| 2022/6/16 | 6 | 5.5 | 2022/7/24 | 694 | 787.8 |
| 2022/6/17 | 7 | 6.5 | 2022/7/25 | 809 | 852.2 |
| 2022/6/18 | 7 | 7.8 | 2022/7/26 | 865 | 921.4 |
| 2022/6/19 | 8 | 9.2 | 2022/7/27 | 977 | 995.4 |
| 2022/6/20 | 8 | 10.9 | 2022/7/28 | 1066 | 1075 |
| 2022/6/21 | 9 | 13 | 2022/7/29 | 1259 | 1160.5 |
| 2022/6/22 | 11 | 15.3 | 2022/7/30 | 1343 | 1251.8 |
| 2022/6/23 | 16 | 18.1 | 2022/7/31 | 1370 | 1350.1 |
| 2022/6/24 | 17 | 21.3 | 2022/8/1 | 1475 | 1455.8 |
| 2022/6/25 | 19 | 25.1 | 2022/8/2 | 1603 | 1568.6 |
| 2022/6/26 | 20 | 29.5 | 2022/8/3 | 1721 | 1689.9 |
| 2022/6/27 | 20 | 34.7 | 2022/8/4 | 1860 | 1820.3 |
| 2022/6/28 | 21 | 40.8 | 2022/8/5 | 2004 | 1959.9 |
| 2022/6/29 | 37 | 48 | 2022/8/6 | 2108 | 2109.7 |
| 2022/6/30 | 49 | 56.4 | 2022/8/7 | 2131 | 2214.2 |
| 2022/7/1 | 64 | 66.2 | 2022/8/8 | 2293 | 2320.1 |
| 2022/7/2 | 76 | 77.7 | 2022/8/9 | 2415 | 2426.9 |
| 2022/7/3 | 78 | 91.2 | 2022/8/10 | 2458 | 2534.8 |
| 2022/7/4 | 80 | 107 | 2022/8/11 | 2458 | 2643.6 |
| 2022/7/5 | 106 | 125.5 | 2022/8/12 | 2746 | 2753.5 |
| 2022/7/6 | 142 | 147.2 | 2022/8/13 | 2848 | 2864.3 |
| 2022/7/7 | 172 | 172.7 | 2022/8/14 | 2893 | 2976.2 |
| 2022/7/8 | 204 | 191.9 | 2022/8/15 | 2985 | 3089.1 |
| 2022/7/9 | 218 | 212.7 | 2022/8/16 | 3183 | 3203 |
| 2022/7/10 | 218 | 235 | 2022/8/17 | 3359 | 3317.9 |
| 2022/7/11 | 228 | 259 | 2022/8/18 | 3450 | 3433.8 |
| 2022/7/12 | 266 | 284.7 | 2022/8/19 | 3655 | 3550.8 |
| 2022/7/13 | 308 | 312.3 | 2022/8/20 | 3755 | 3668.7 |
| 2022/7/14 | 347 | 342 | 2022/8/21 | 3787 | 3787.7 |
| 2022/7/15 | 347 | 373.7 | 2022/8/22 | 3896 | 3907.7 |

| Date | Observed cumulated cases | Simulated cumulated cases | Date | Observed cumulated cases | Simulated cumulated cases |
|---|---|---|---|---|---|
| 2022/8/23 | 3984 | 4028.9 | | | |
| 2022/8/24 | 4144 | 4151.2 | | | |
| 2022/8/25 | 4216 | 4274.6 | | | |
| 2022/8/26 | 4472 | 4399.1 | | | |
| 2022/8/27 | 4472 | 4524.6 | | | |
| 2022/8/28 | 4472 | 4651.3 | | | |
| 2022/8/29 | 4692 | 4779 | | | |
| 2022/8/30 | 4876 | 4907.9 | | | |
| 2022/8/31 | 5037 | 5037.8 | | | |
| 2022/9/1 | 5197 | 5150.4 | | | |
| 2022/9/2 | 5197 | 5259.4 | | | |
| 2022/9/3 | 5197 | 5367.4 | | | |
| 2022/9/4 | 5409 | 5474.2 | | | |
| 2022/9/5 | 5525 | 5579.9 | | | |
| 2022/9/6 | 5692 | 5684.4 | | | |
| 2022/9/7 | 5726 | 5787.9 | | | |
| 2022/9/8 | 5852 | 5890.2 | | | |
| 2022/9/9 | 5971 | 5991.4 | | | |
| 2022/9/10 | 6014 | 6091.6 | | | |
| 2022/9/11 | 6032 | 6190.7 | | | |
| 2022/9/12 | 6129 | 6288.7 | | | |
| 2022/9/13 | 6246 | 6385.8 | | | |
| 2022/9/14 | 6448 | 6481.9 | | | |
| 2022/9/15 | 6649 | 6576.9 | | | |
| 2022/9/16 | 6806 | 6670.9 | | | |
| 2022/9/17 | 6867 | 6763.9 | | | |
| 2022/9/18 | 6867 | 6855.9 | | | |
| 2022/9/19 | 7018 | 6946.9 | | | |
| 2022/9/20 | 7115 | 7036.9 | | | |
| 2022/9/21 | 7205 | 7125.8 | | | |
| 2022/9/22 | 7300 | 7213.9 | | | |
| 2022/9/23 | 7300 | 7301 | | | |

### (e) United Kingdom

| Date | Observed cumulated cases | Simulated cumulated cases | Date | Observed cumulated cases | Simulated cumulated cases |
|---|---|---|---|---|---|
| 2022/5/6 | 1 | 1.0 | 2022/6/13 | 470 | 445.4 |
| 2022/5/7 | 1 | 1.3 | 2022/6/14 | 524 | 474.6 |
| 2022/5/8 | 1 | 1.7 | 2022/6/15 | 524 | 504.8 |
| 2022/5/9 | 1 | 2.1 | 2022/6/16 | 574 | 536.2 |
| 2022/5/10 | 1 | 2.6 | 2022/6/17 | 574 | 568.9 |
| 2022/5/11 | 1 | 3.2 | 2022/6/18 | 574 | 602.7 |
| 2022/5/12 | 2 | 4.0 | 2022/6/19 | 574 | 637.7 |
| 2022/5/13 | 3 | 4.8 | 2022/6/20 | 793 | 674.0 |
| 2022/5/14 | 3 | 5.8 | 2022/6/21 | 793 | 711.7 |
| 2022/5/15 | 7 | 7.0 | 2022/6/22 | 793 | 750.9 |
| 2022/5/16 | 7 | 8.5 | 2022/6/23 | 910 | 791.6 |
| 2022/5/17 | 7 | 10.2 | 2022/6/24 | 910 | 833.6 |
| 2022/5/18 | 9 | 12.2 | 2022/6/25 | 910 | 877.2 |
| 2022/5/19 | 9 | 14.5 | 2022/6/26 | 1076 | 922.5 |
| 2022/5/20 | 20 | 17.3 | 2022/6/27 | 1076 | 969.6 |
| 2022/5/21 | 20 | 20.6 | 2022/6/28 | 1076 | 1018.4 |
| 2022/5/22 | 20 | 24.6 | 2022/6/29 | 1076 | 1069.0 |
| 2022/5/23 | 57 | 29.2 | 2022/6/30 | 1235 | 1121.3 |
| 2022/5/24 | 71 | 34.7 | 2022/7/1 | 1235 | 1175.6 |
| 2022/5/25 | 78 | 41.2 | 2022/7/2 | 1235 | 1232.0 |
| 2022/5/26 | 106 | 48.9 | 2022/7/3 | 1235 | 1290.4 |
| 2022/5/27 | 106 | 58.0 | 2022/7/4 | 1351 | 1351.4 |
| 2022/5/28 | 106 | 68.8 | 2022/7/5 | 1351 | 1395.5 |
| 2022/5/29 | 106 | 81.6 | 2022/7/6 | 1351 | 1440.5 |
| 2022/5/30 | 179 | 96.8 | 2022/7/7 | 1552 | 1485.2 |
| 2022/5/31 | 190 | 114.7 | 2022/7/8 | 1552 | 1529.8 |
| 2022/6/1 | 196 | 135.9 | 2022/7/9 | 1552 | 1574.2 |
| 2022/6/2 | 207 | 161.0 | 2022/7/10 | 1552 | 1618.3 |
| 2022/6/3 | 226 | 190.7 | 2022/7/11 | 1735 | 1662.3 |
| 2022/6/4 | 226 | 226.0 | 2022/7/12 | 1735 | 1706.1 |
| 2022/6/5 | 226 | 246.8 | 2022/7/13 | 1735 | 1749.7 |
| 2022/6/6 | 302 | 268.5 | 2022/7/14 | 1856 | 1793.0 |
| 2022/6/7 | 321 | 291.1 | 2022/7/15 | 1856 | 1836.2 |
| 2022/6/8 | 321 | 314.6 | 2022/7/16 | 1856 | 1879.3 |
| 2022/6/9 | 366 | 338.8 | 2022/7/17 | 1856 | 1922.1 |
| 2022/6/10 | 366 | 364.0 | 2022/7/18 | 2137 | 1964.7 |
| 2022/6/11 | 366 | 390.1 | 2022/7/19 | 2137 | 2007.1 |
| 2022/6/12 | 470 | 417.3 | 2022/7/20 | 2137 | 2049.3 |

| Date | Observed cumulated cases | Simulated cumulated cases | Date | Observed cumulated cases | Simulated cumulated cases |
|---|---|---|---|---|---|
| 2022/7/21 | 2208 | 2091.4 | 2022/8/28 | 3340 | 3428.3 |
| 2022/7/22 | 2208 | 2133.2 | 2022/8/29 | 3413 | 3440.0 |
| 2022/7/23 | 2208 | 2174.9 | 2022/8/30 | 3413 | 3451.0 |
| 2022/7/24 | 2208 | 2216.3 | 2022/8/31 | 3413 | 3461.4 |
| 2022/7/25 | 2497 | 2257.6 | 2022/9/1 | 3413 | 3471.2 |
| 2022/7/26 | 2497 | 2298.7 | 2022/9/2 | 3413 | 3480.3 |
| 2022/7/27 | 2497 | 2339.7 | 2022/9/3 | 3413 | 3488.9 |
| 2022/7/28 | 2546 | 2380.4 | 2022/9/4 | 3413 | 3497.0 |
| 2022/7/29 | 2546 | 2420.9 | 2022/9/5 | 3484 | 3504.6 |
| 2022/7/30 | 2546 | 2461.3 | 2022/9/6 | 3484 | 3511.7 |
| 2022/7/31 | 2546 | 2501.5 | 2022/9/7 | 3484 | 3518.4 |
| 2022/8/1 | 2759 | 2541.5 | 2022/9/8 | 3484 | 3524.7 |
| 2022/8/2 | 2759 | 2581.3 | 2022/9/9 | 3484 | 3530.6 |
| 2022/8/3 | 2759 | 2620.9 | 2022/9/10 | 3484 | 3536.2 |
| 2022/8/4 | 2859 | 2660.4 | 2022/9/11 | 3484 | 3541.4 |
| 2022/8/5 | 2859 | 2699.7 | 2022/9/12 | 3552 | 3546.3 |
| 2022/8/6 | 2859 | 2738.7 | 2022/9/13 | 3552 | 3550.9 |
| 2022/8/7 | 2859 | 2777.6 | 2022/9/14 | 3552 | 3555.2 |
| 2022/8/8 | 3017 | 2816.4 | 2022/9/15 | 3552 | 3559.2 |
| 2022/8/9 | 3017 | 2854.9 | 2022/9/16 | 3552 | 3563.0 |
| 2022/8/10 | 3017 | 2893.3 | 2022/9/17 | 3552 | 3566.6 |
| 2022/8/11 | 3017 | 2931.5 | 2022/9/18 | 3552 | 3570.0 |
| 2022/8/12 | 3017 | 2969.5 | 2022/9/19 | 3552 | 3573.1 |
| 2022/8/13 | 3017 | 3007.4 | 2022/9/20 | 3585 | 3576.1 |
| 2022/8/14 | 3017 | 3045.1 | 2022/9/21 | 3585 | 3578.9 |
| 2022/8/15 | 3195 | 3082.6 | 2022/9/22 | 3585 | 3581.5 |
| 2022/8/16 | 3195 | 3119.9 | 2022/9/23 | 3585 | 3583.9 |
| 2022/8/17 | 3195 | 3157.1 | | | |
| 2022/8/18 | 3195 | 3194.1 | | | |
| 2022/8/19 | 3195 | 3230.9 | | | |
| 2022/8/20 | 3195 | 3267.5 | | | |
| 2022/8/21 | 3195 | 3304.0 | | | |
| 2022/8/22 | 3340 | 3340.6 | | | |
| 2022/8/23 | 3340 | 3357.0 | | | |
| 2022/8/24 | 3340 | 3373.1 | | | |
| 2022/8/25 | 3340 | 3388.1 | | | |
| 2022/8/26 | 3340 | 3402.3 | | | |
| 2022/8/27 | 3340 | 3415.7 | | | |

**Figure A1. The observed cumulative Monkeypox cases and predicted cumulated Monkeypox cases on the global and in the different countries by the deterministic SIR model.**

(a) The global

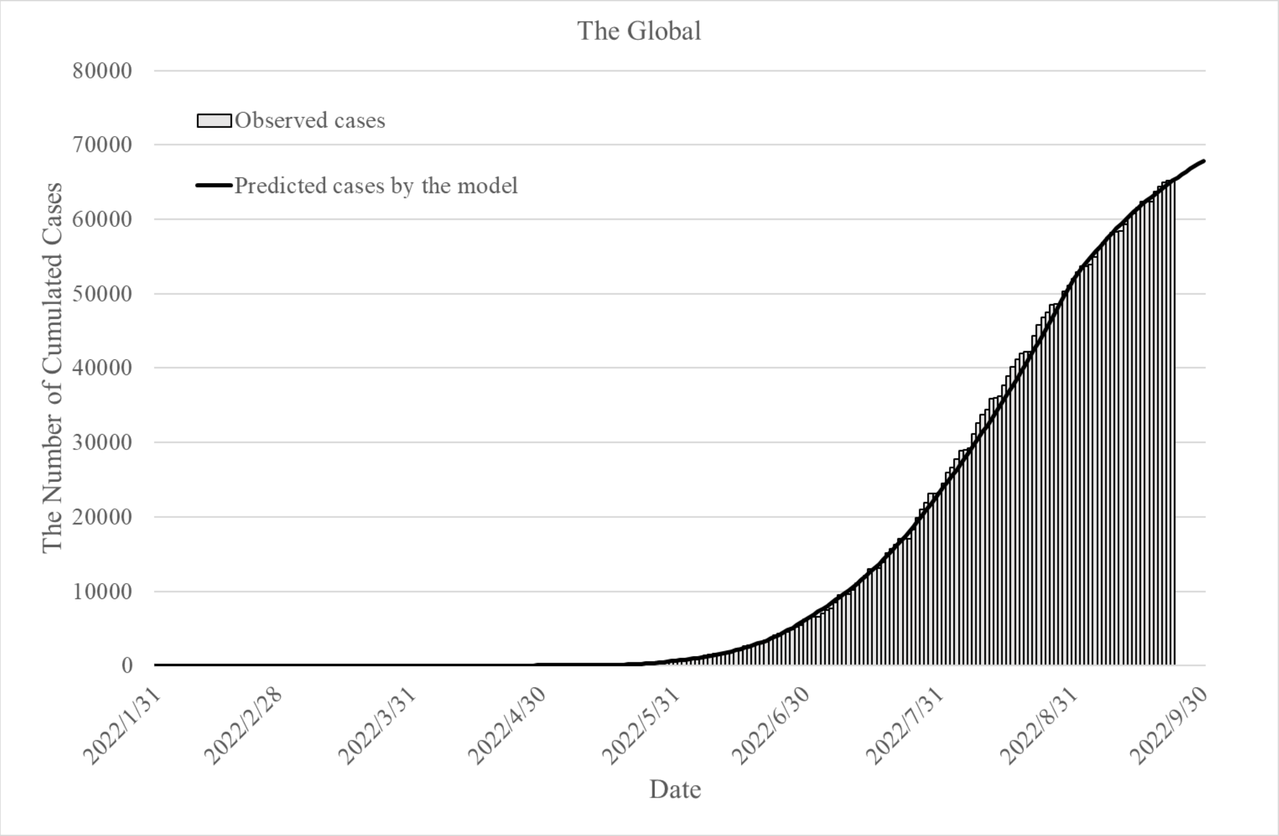

(b) United States

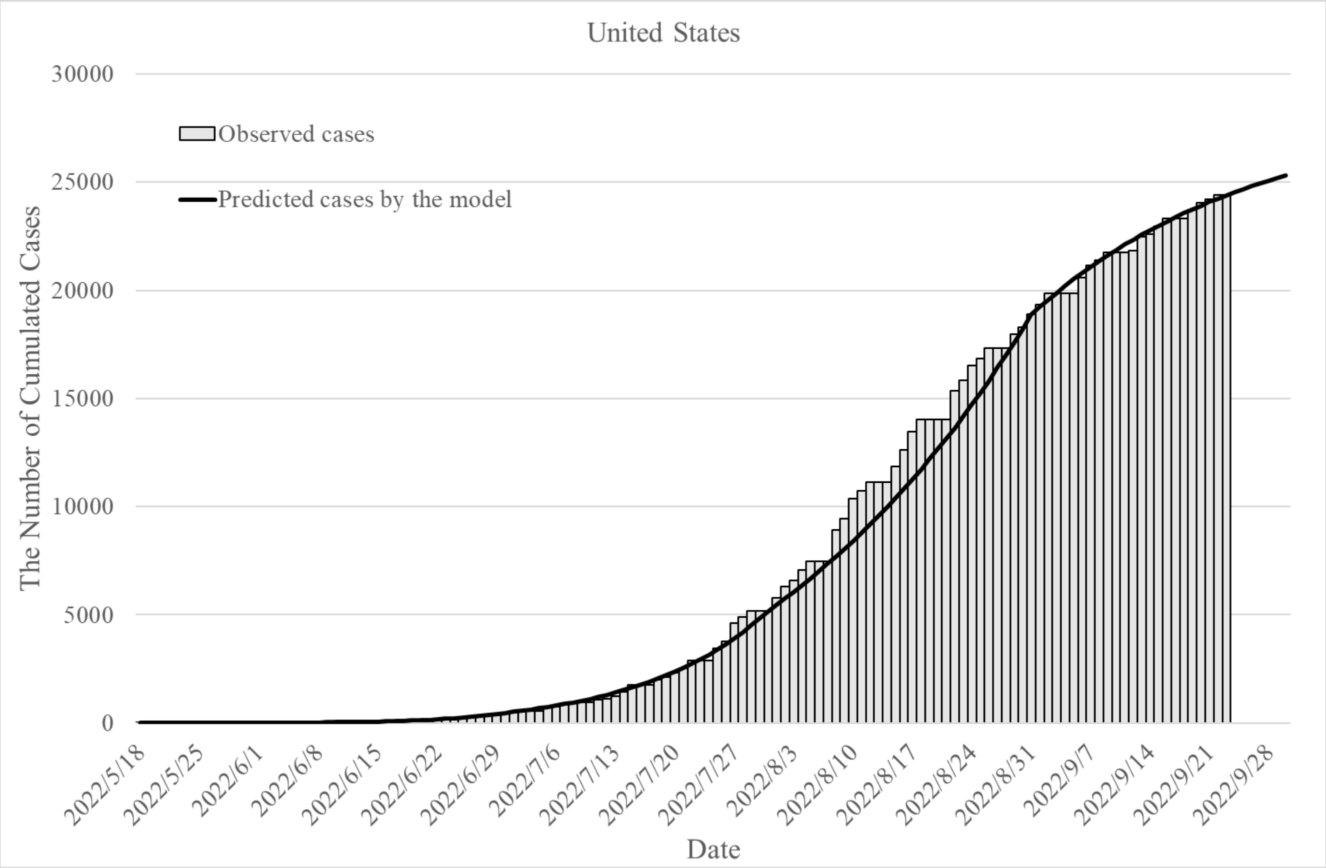

(c) Spain

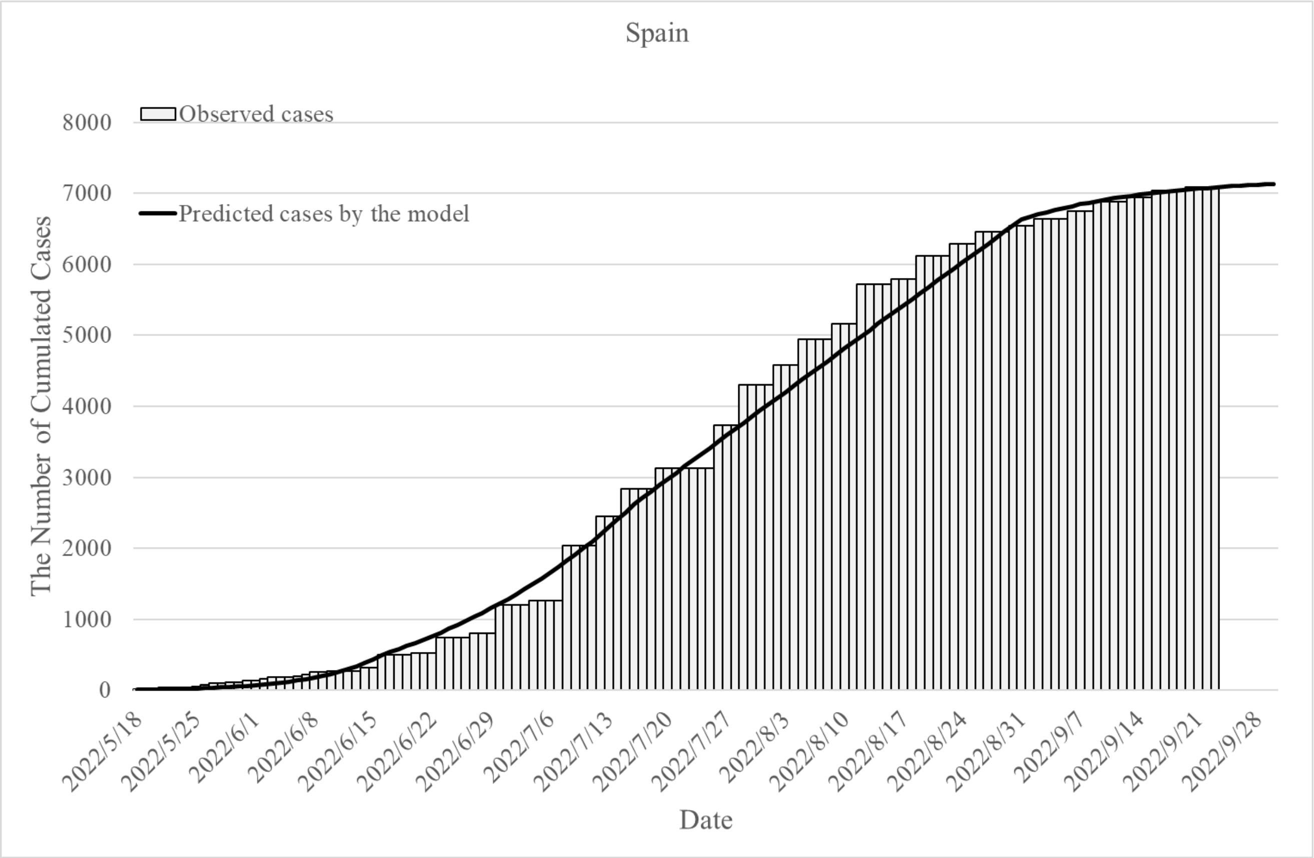

Spain
Observed cases
Predicted cases by the model
8000
7000
6000
5000
4000
3000
2000
1000
0
The Number of Cumulated Cases
2022/5/18
2022/5/25
2022/6/1
2022/6/8
2022/6/15
2022/6/22
2022/6/29
2022/7/6
2022/7/13
2022/7/20
2022/7/27
2022/8/3
2022/8/10
2022/8/17
2022/8/24
2022/8/31
2022/9/7
2022/9/14
2022/9/21
2022/9/28
Date

(d) Brazil

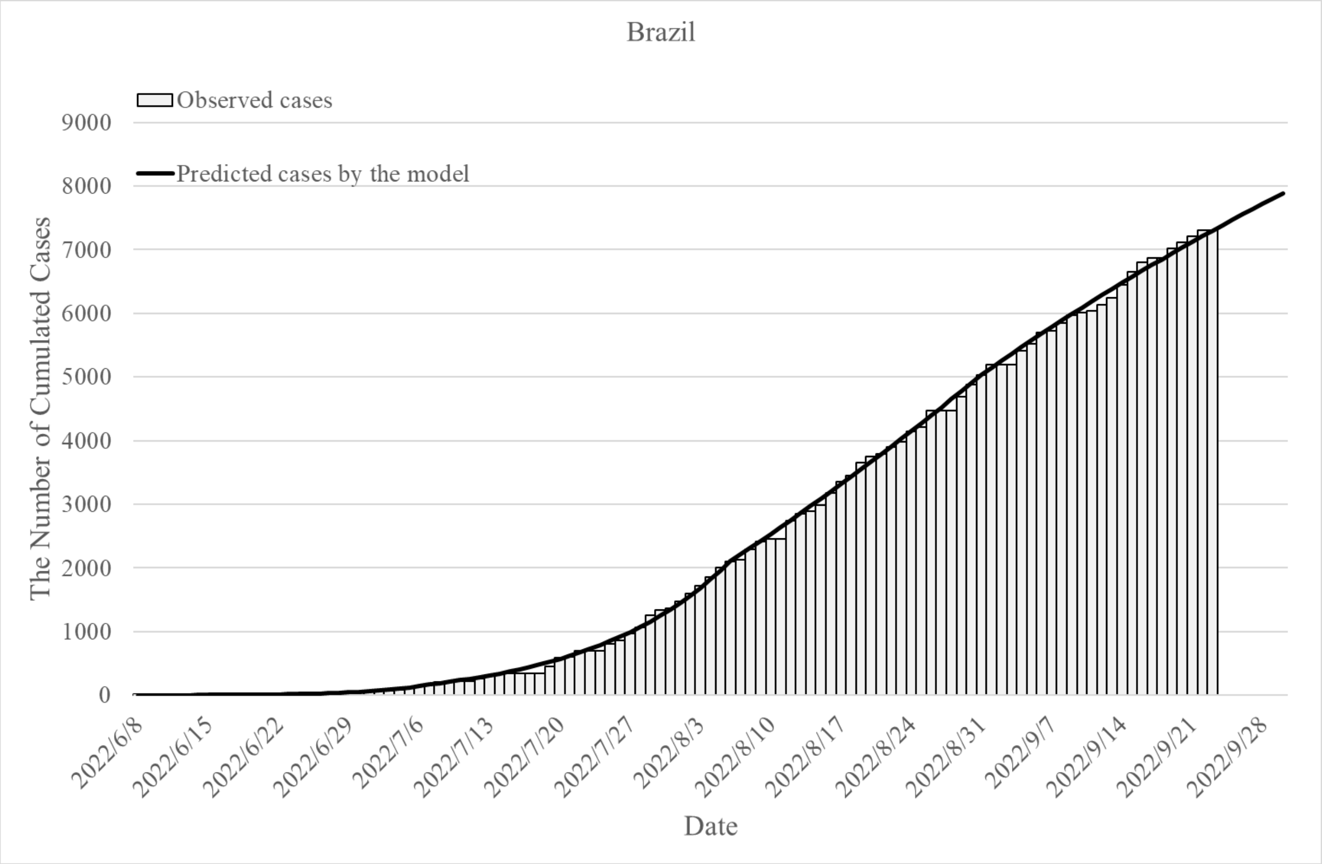

Brazil
Observed cases
Predicted cases by the model
9000
8000
7000
6000
5000
4000
3000
2000
1000
0
The Number of Cumulated Cases
2022/6/8
2022/6/15
2022/6/22
2022/6/29
2022/7/6
2022/7/13
2022/7/20
2022/7/27
2022/8/3
2022/8/10
2022/8/17
2022/8/24
2022/8/31
2022/9/7
2022/9/14
2022/9/21
2022/9/28
Date

(e) United Kingdom

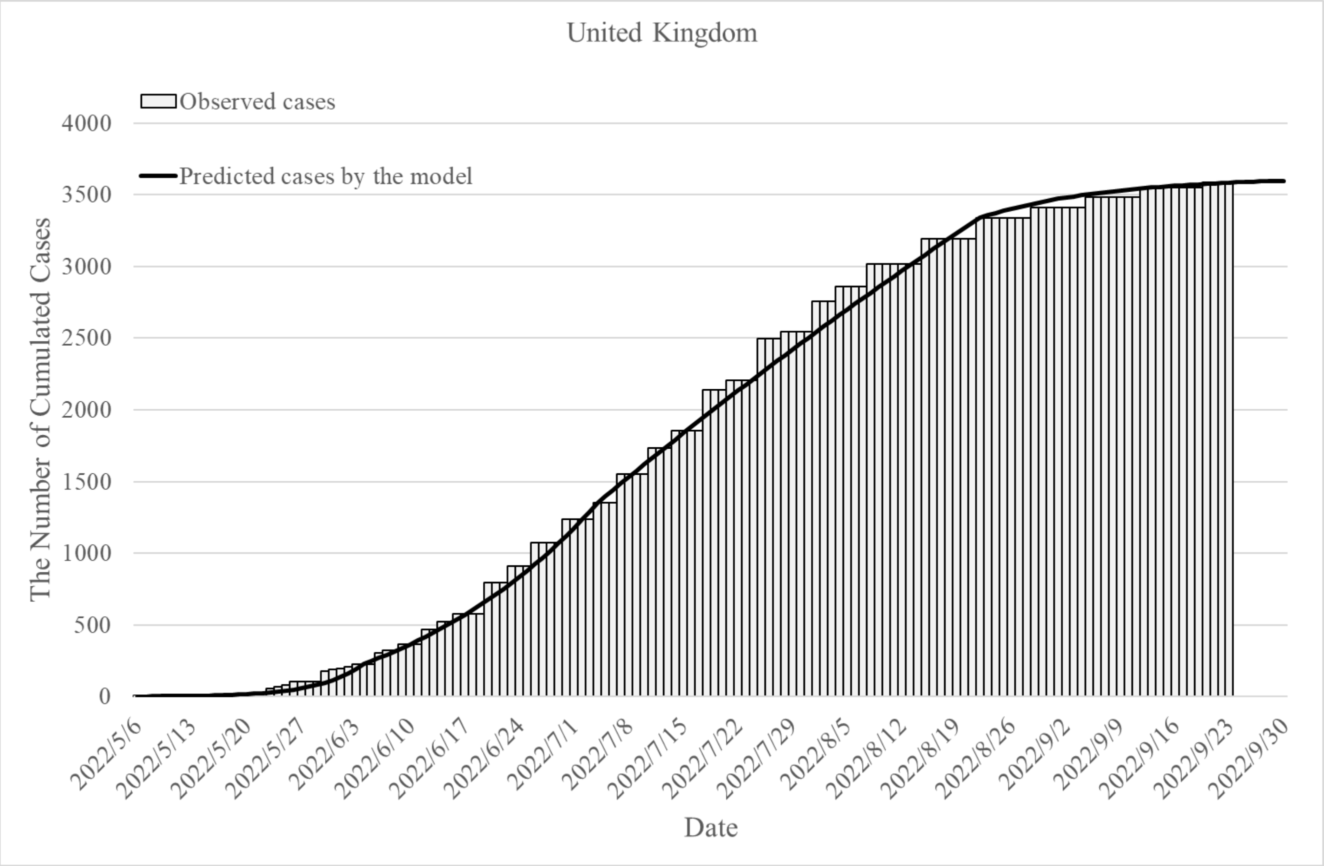
